# Protocol for an adaptive platform trial of intended service user-derived interventions to equitably reduce non-attendance in eye screening programmes in Botswana, India, Kenya & Nepal

**DOI:** 10.1101/2024.07.16.24310491

**Authors:** Luke Allen, Min Kim, Malebogo Tlhajoane, David Macleod, Oathokwa Nkomazana, Michael Gichangi, Sailesh Kumar Mishra, Shalinder Sabherwal, James Carpenter, Sarah Karanja, Ari Ho-Foster, Bakgaki Ratshaa, Nigel Bolster, Jacqui Ramke, Matthew Burton, Andrew Bastawrous

**Affiliations:** LSHTM; University of Botswana; LSHTM and Kenyan Ministry of Health; LSHTM and UCL; KEMRI, Kenya

**Keywords:** Health services research, platform trial, embedded trial, global health, mHealth, equity

## Abstract

**Background:** Only 30-50% of people referred to clinics during community-based eye screening are able to access care in Botswana, India, Kenya, and Nepal. The access rate is even lower for certain population groups. This platform trial aims to test multiple, iterative, low-risk public health interventions and simple service modifications with a series of individual randomised controlled trials (RCT) conducted in each country, with the aim of increasing the proportion of people attending.

**Methods and Analysis:** We will set up a platform trial in each country to govern the running of a series of pragmatic, adaptive, embedded, parallel, multi-arm, superiority RCTs to test a series of service modifications suggested by intended service users. The aim is to identify serial marginal gains that cumulatively result in large improvements to equity and access. The primary outcome will be the probability of accessing treatment among the population group with the worst access at baseline. We will calculate Bayesian posterior probabilities of clinic attendance in each arm every 72 hours. Each RCT will continually recruit participants until the following default stopping rules have been met: >95% probability that one arm is best; >95% probability that the difference between the best arm and the arms remaining in the trial is <1%; or 10,000 people have been recruited. Lower thresholds may be used for RCTs testing interventions with very low risks and costs. The specific design of cluster RCTs will be determined by our research team once the intervention is known, but the population and outcome will be the same across all trials.

**Discussion:** This APT will be used to identify effective service modifications, driving continuous improvements in access.

**Ethics and Dissemination:** This trial has been approved by the research ethics committee at the London School of Hygiene and Tropical Medicine. Approvals for individual interventions will be sought from UK and local ethics committees. Results will be shared via local workshops, social media, and peer-reviewed publications.

**Trial Registration:** ISRCTN <u>53970958</u>. Registered on 21 September 2023

**Strengths and Limitations:**

- Randomised control trials are resource intensive and often require lengthy set up periods. The adaptive platform design allows for the evaluation of multiple interventions with a single outcome, governed by a predefined set of criteria
- The study defaults are designed to test multiple low-risk, incremental service modifications in series, and quickly identify those that are just as good as, or superior to the status quo.
- Our high default tolerance for type I error means that we will often incorrectly identify arms as superior when really there is no difference. This is acceptable when arms confer similar costs and negligible risks.
- Our default very low type II error rate means that we will very rarely mistakenly identify an inferior arm as being superior.
- Our trial is embedded within screening programmes and uses automated randomisation, allocation, data collection, and statistical testing to minimise resource requirements.

## Introduction

This protocol sets out the approach for running platform trials in four countries that will test interventions suggested by local intended service beneficiaries with the intention of improving equitable access to community-based eye services. Box 1 sets out the definitions of common terms used in the protocol.

Many health programmes experience large mismatches between those identified with a clinical need and those who access services. A recent international systematic review of ‘no-show’ appointments across all medical specialities in primary and secondary care estimated that 23% of clinic appointments are not attended, with the highest rate observed on the African continent (43%).^1^ Complex supply and demand factors govern access to health services,^2^ and systematically marginalised populations are often the least likely to receive care.^3,4^ Improving access to care lies at the heart of Universal Health Coverage (UHC) and is a core element in the Sustainable Development Agenda.^5^

Eye services offer an instructive case study. Approximately 1.1 billion people (over 10% of the global population) live with vision impairment that could be corrected.^6^ Two very cost-effective interventions - spectacles and cataract surgery – could eliminate over 90% of all vision impairment worldwide. Although provision of these services has risen in recent decades, effective coverage rates exhibit marked socioeconomic gradients at the international and intra-national levels, for example, the global effective refractive coverage is reported at 36%, with high-income countries reporting 90% and low-income only 6%.^6^

In major eye screening programmes, once people have been identified with an eye need and referred, typically only around 30-50% of these people receive care. Often there are marked socioeconomic inequalities in terms of which groups face the highest barriers to eye care.^6–8^

#### Box 1: Terms used in this protocol

##### Access and attendance

We are interested in **access to services**, which is driven by complex supply and demand factors. We will use **attendance** as the primary indicator of access, but note that this term can carry the implication that intended service beneficiaries are to blame when in reality, it is often features of the service that present unsurmountable obstacles to access, especially for left behind groups. We also note that both access and attendance are proximal outcomes, in that they do not automatically lead to the receipt of good quality care and improved health outcomes.

##### Eye care need

We are concerned with whether those with an **eye care need** access services. This includes near or distance vision impairment and non-visually impairing eye conditions, included but not limited to: uncorrected and under-corrected refractive errors, cataract, eye redness, eye discomfort or pain, or any other eye-related issue identified by screeners.

##### Left behind population groups

We focus on the **population groups** with the worst access to services, aligning with the UN Agenda for Sustainable Development’s “central, transformative promise” to ‘leave no one behind’ and ‘reach the furthest behind first’. Further UN guidance states that “leaving no one behind means moving beyond assessing average and aggregate progress, towards ensuring progress for all population groups at a disaggregated level.” The UN uses the terms ‘worst-off’ and ‘left-behind’ groups interchangeably. Multiple population subgroups and domains can be used for disaggregation including as age, sex, ethnicity, occupation, income, socioeconomic status etc.

##### Platform trial

Platform trials use shared infrastructure and a master protocol to run multiple individual trials that test different interventions against a constant outcome (attendance) in the same target population (people identified with an eye need at screening and referred for further care).

##### Individual trial

A randomised controlled trial of a single intervention (e.g. a voucher or SMS reminder message) that is performed under the platform trial protocol.

##### Intervention/service modification

We use the term **‘intervention’** when a new element is added to programmes (such as vouchers), and **‘service modification’** when an existing element is tweaked, such as amending opening hours, or the wording used in communication materials.

##### Arms

Variants or ‘doses’ of the intervention/service modification. These are tested against each other and a control arm. For instance, an individual trial might test vouchers (the intervention) with three different arms; $1, $5, and $10 against a control arm (no voucher).

##### Embedded

The trial takes place within a real-world clinical programme, using routinely collected data.

##### Pragmatic

The interventions will be delivered to typical patients by programme staff (rather than research staff).

##### Adaptive

The algorithm will use of stopping rules to run regular interim analyses. Recruitment will continue until one or more of the stopping rules is met, meaning that sample size will be optimised.

##### Bayesian

The testing algorithm will use a Bayesian rather than a frequentist statistical approach; incorporating prior beliefs into the analysis and then using accruing data to continuously update the probability of events, as probability distributions.

Our research collaborative (LSHTM, Peek Vision, COESCA, Kenyan MoH, University of Botswana, NNJS, Dr Shroff’s Charity Eye Hospital) is working with four major eye screening programmes to identify the population groups least able to access care in each setting (Table 1).

**Table 1.** Eye screening programmes.

| Country | Programme description | Dates | Population |
| --- | --- | --- | --- |
| Botswana | The 'Pono Yame' national school-based programme. Screeners travel to every school in the country and refer positive cases to local triage and treatment camps | 2022-2024 | One national programme: 500,000 children aged 5-18y |
| India | House-to-house community-based screening in three sites in central Uttar Pradesh. | 2023-2025 | Three sites: each with 50,000 to 70,000 adults and children. |
| Kenya | Community-based screening programmes in Meru and Kwale with school-based and primary care facility-based screening. | 2022-2025 | Two sites with: each with approximately 1 million adults and children |
| Nepal | Regional primary care-based passive screening programme in Rajbiraj, Eastern Nepal. | 2022-2023 | One regional site with approximately 70,000 adults and children |

Through interviewing representatives from the sociodemographic groups that face the highest barriers to care, we aim to identify potential service modifications that could equitably improve access. Testing whether these intended service user-derived service modifications are causally associated with positive change requires the use of randomisation.

Randomised control trials (RCTs) provide the most robust means of appraising whether an intervention is causally associated with a change in a given outcome. Unfortunately, the resources and technical expertise required to run an RCT generally preclude their use by day-to-day health services. To overcome this barrier, we are proposing use of an automated RCT platform embedded within app-based patient workflow screening and referral systems to perform elements of randomisation, allocation, outcome assessment, and statistical testing. Global health programmes constantly adapt in order to better meet the needs of their beneficiaries, however the impact of these adaptations is rarely assessed. By reducing the barriers for rigorously testing service modifications, we hope to equip programme managers with the ability to run resource-light, real-time, embedded RCTs to continuously improve their programmes and address socioeconomic inequalities in attendance and outcomes.

Rather than running serial RCTs – each requiring lengthy set-up periods and very similar protocols, we intend to set up a platform trial. This design uses a master protocol to evaluate multiple interventions in the context of a single outcome in a perpetual manner. Platform trials are a form of multi-intervention, multi-stage design.^10^ Figure 1 illustrates how multiple different interventions can be tested in individual trials under a single overarching platform trial protocol.

**Figure 1:**
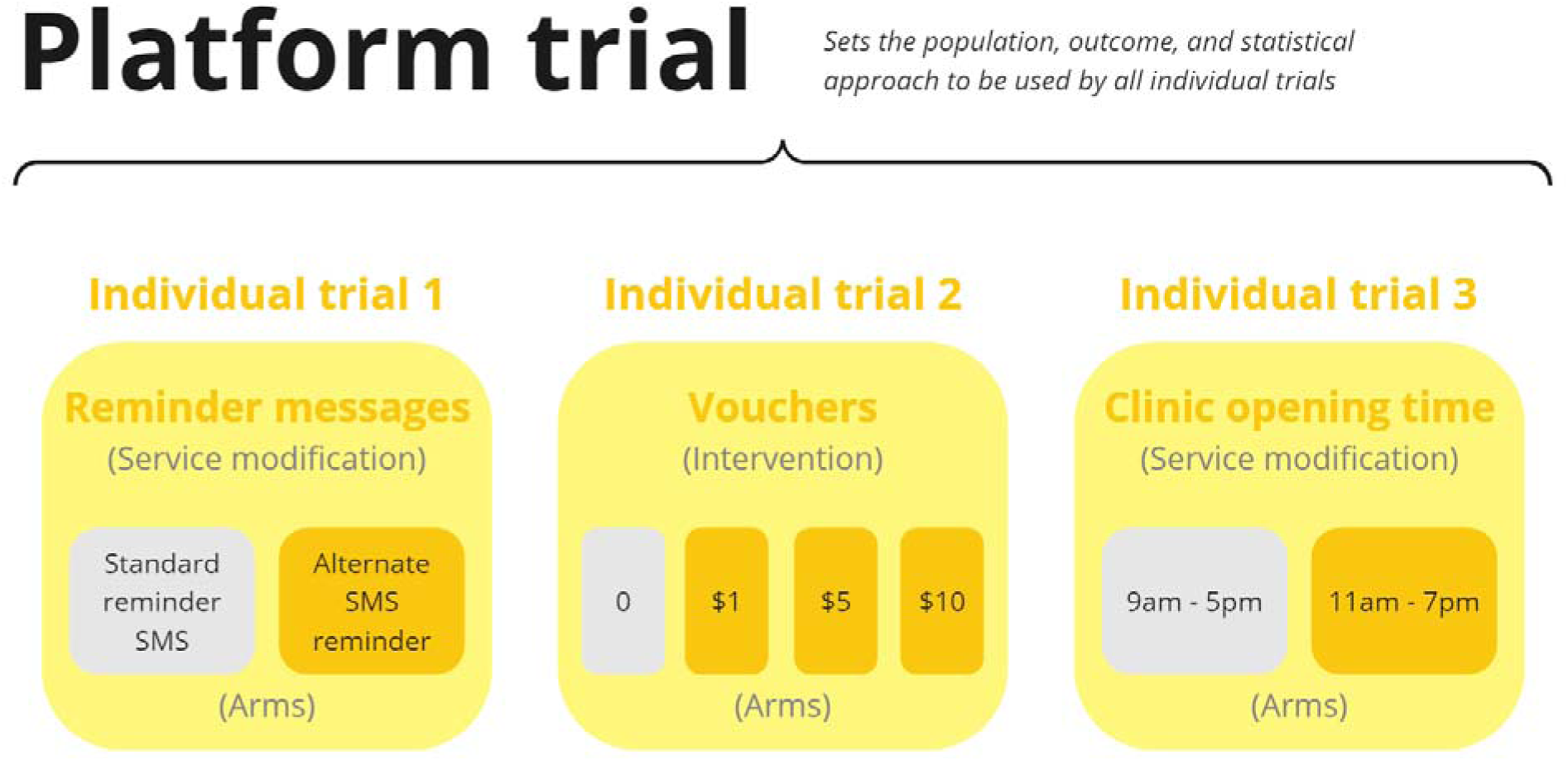
Three example individual trials that test new interventions against the status quo (grey boxes) as part of an overall platform trial

Addressing inequitable service outcomes is likely to require multiple different modifications in the context of continuous improvement. Early data from Botswana suggests that approximately 1/10 schoolchildren have an unmet eye health need but less than a third are able to access community eye clinics to receive care. Data from Kenya suggests that only a third of younger adults identified with an eye need are able to access care.

In each setting where Peek eye screening programmes run, we intend to engage with representatives from groups that are facing the highest barriers to accessing care to explore their perceptions of the types of interventions and service modifications that could improve access. Our platform trial will be used to test the interventions suggested by these left behind groups.

### Objectives

This platform trial will iteratively test a series of interventions selected with intended service beneficiaries to increase attendance rates in community-based eye screening programmes in Botswana, India, Kenya and Nepal. Each of these programmes use the mature and validated app-based screening system developed by Peek Vision.^11–15^ Programme managers in each country are interested in identifying incremental gains from multiple service modifications to deliver iterative improvements in equitable access.

### Trial design

This Bayesian, embedded, pragmatic, superiority, platform trial protocol will be used to evaluate multiple interventions against a control group, using a constant outcome. The same platform approach will be used in each setting, but the interventions will all be locally-derived and tested. In each setting, the platform trial will be embedded into the local eye screening programme, using referral and attendance data directly derived from the patient management and flow software in each setting.

### Study setting

Platform trials will be established in regional and national community-based eye screening programmes in Botswana (national), Nepal (one regional site), Kenya (two regional sites), and India (three regional sites). All seven sites operate using integrated screening and patient management software developed by Peek Vision. In each site our platform trial will use data routinely gathered using Peek software.

Peek Vision is a leading provider of eye screening software worldwide. The ‘Peek Capture’ app is used to screen participants for vision impairment, to capture observations by screeners and health practitioners, and to gather demographic data, as well as linking participants to a referral system that tracks each of their progression through the local eye health system. The same app is used to collect data on visual acuity, socioeconomic status, referral status, and attendance status (our primary outcome). Our trial will use these routinely collected data to test whether a series of interventions are able to reduce the proportion of people from marginalised groups with an eye care need who do not attend triage clinic once referred (Figure 2).

**Figure 2:**
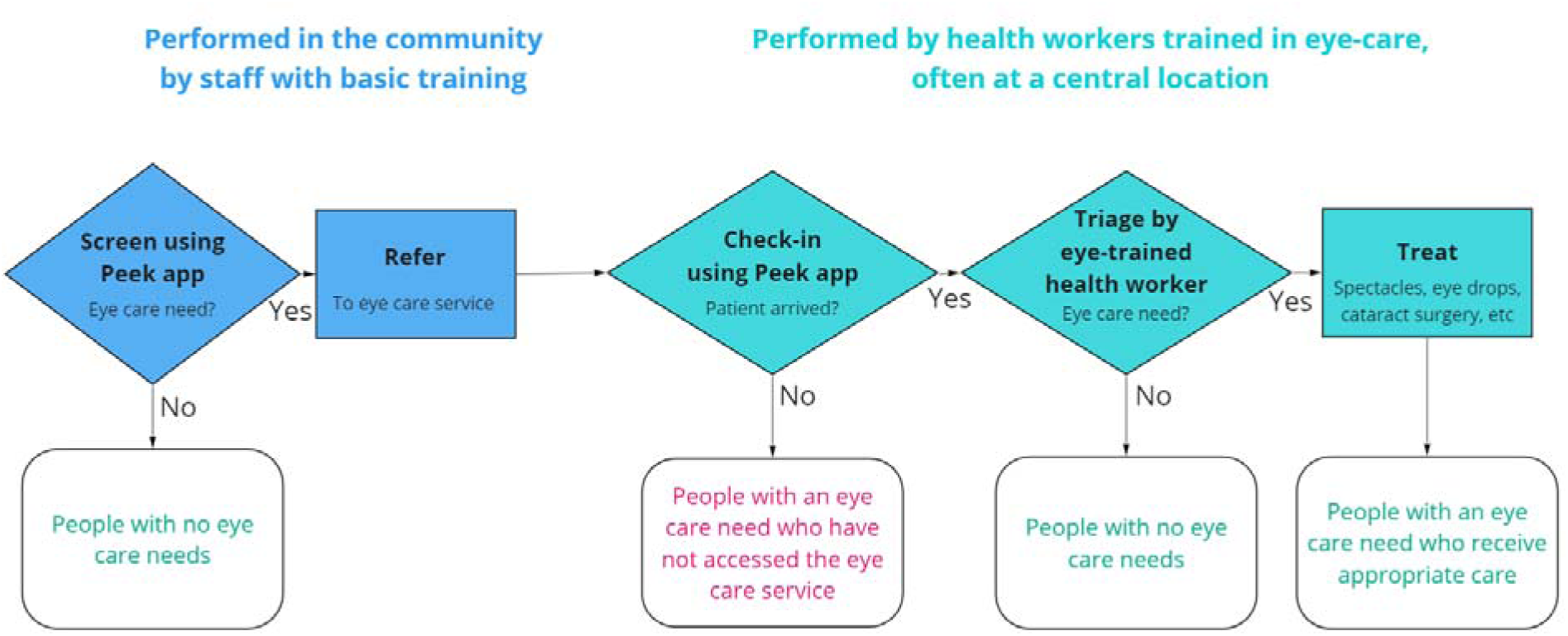
Schematic of patient flow through a Peek programme

### Eligibility criteria

As a pragmatic trial, the eligibility criteria are determined by local programmes. We will include children aged over 5 years and adults who participate in Peek-powered eye screening programmes as outlined in Table 2. We will exclude those who do not meet local clinical service eligibility criteria, such as age (most programmes exclude children younger than 5 years).

**Table 2:** Examples of digital and non-digital interventions delivered at the individual and cluster levels.

| Type of intervention | Individual | Cluster |
| --- | --- | --- |
| **Digital** | No human input required for intervention delivery. Peek software performs random allocation and delivers the intervention e.g.<br>• SMS messages<br>• Pre-recorded voice messages<br>• Visual acuity thresholds<br>• *eVouchers* | Humans select the clusters and the Peek software delivers interventions e.g.<br>• SMS messages sent to a headteacher<br>• Messages sent to a village chief<br>• Pre-recorded voice messages sent to a primary care facility manager |
| **Non-digital** | Peek software performs random allocation then informs the human team. They deliver the interventions e.g.<br>• Physical vouchers<br>• Chaperones<br>• Individualised transport assistance | Peek software can randomise the clusters, but humans need to deliver the interventions e.g.<br>• Radio broadcasts<br>• Training for implementers<br>• New clinic times or locations<br>• New bus services |

Note that this trial will not test any pharmaceutical or medical interventions: the focus is on low-risk service modifications and public health interventions.

### Interventions

#### Interventions and administration

This platform trial is being set up to test service modifications suggested by representatives of groups that face the highest barriers to receiving care. The intention is to continuously improve attendance rates with the greatest gains focused on left behind groups.

This platform trial forms the testing element of a broader continuous improvement model called ‘IM-SEEN’ (IMprovement Studies for Equitable and Evidence-based iNnovation). The model has already been integrated into Peek programmes (orange boxes shown in Figure 3). In this continuous improvement approach, data collectors gather contact details and sociodemographic data from those found to have an eye problem prior to referring them. This means that programme managers using Peek have a complete record of who has not attended clinic on the appointed day, and they are able to identify the population group with the lowest attendance. Next, the programme leadership team can engage with representatives of left-behind groups to elicit barriers and identify potential service improvements that would reduce non-attendance – such as changing the clinic location or amending the wording of the SMS reminder messaging. The final step is to use embedded RCTs to test these proposed improvements with new referrals. Effective interventions will be adopted across the programme. Further information on the broader IM-SEEN approach has been published elsewhere.^8^

**Figure 3:**
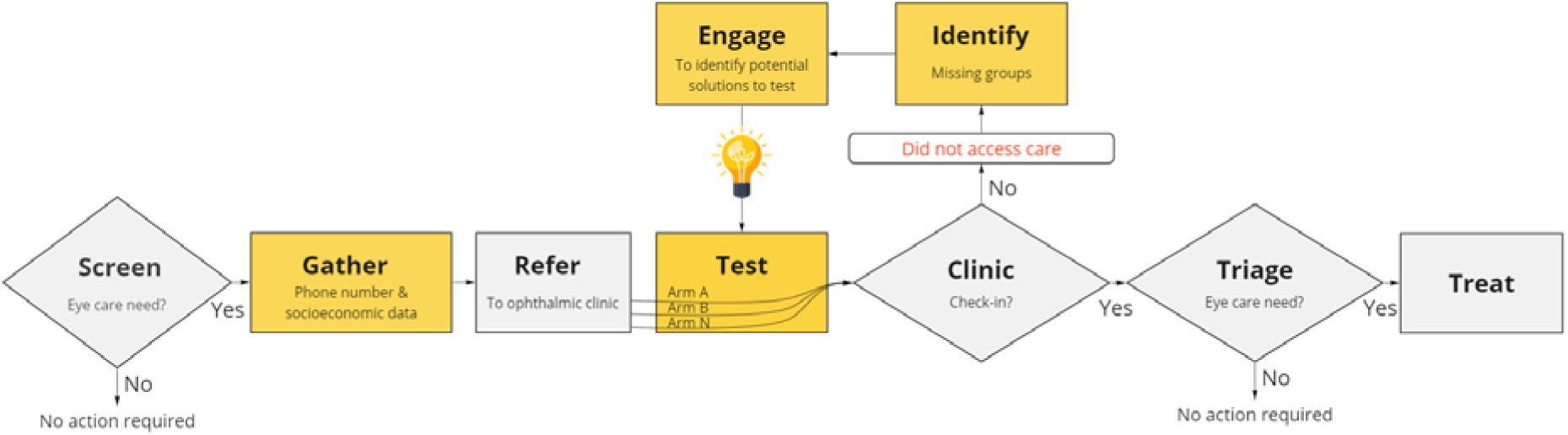
Elements in the IM-SEEN continuous improvement model

Screeners collect data on age, sex, location, language, ethnicity, health status, education, occupation, and income/assets, with minor local variations and enter these data into the Peek app directly after screening. Some of these categories are binary whereas other have multiple response options e.g. language. In all, the survey data can be used to divide screening populations into approximately 60 different groups, each defined by a single characteristic e.g. ‘female’ or ‘primary school education’. We perform multivariable logistic regression to identify which population subgroups have the lowest attendance in each site.

We then conduct interviews with members of the group with the lowest attendance to identify potential service modifications to improve attendance. Rather than designing de-novo interventions, or selecting complex interventions, the focus of this process is on identifying very simple service modifications such as changing the time, day, or location of clinics, changing the language or wording of reminder SMS messages, or providing simple incentives like vouchers. There is scope to identify other ‘off-the-shelf’ interventions that have previously been shown to work in other contexts, but the focus is firmly on translation and implementation research rather than discover or knowledge generation i.e., the platform trial will be used to run ‘T3/T4’ implementation studies in each site.^16^

Once the elicitation studies have generated a list of potential service modifications, a local management group comprised of community representatives, programme managers, public health experts and programme managers (Box 2) will select a shortlist of service modification that can be tested, based on the following criteria:

- **Impact**: is the intervention likely to improve attendance? i.e., has this intervention been tested in other contexts and demonstrated a meaningful effect?
- **Risk**: what level of risk does the intervention pose to service users?
- **Feasibility**: how easily can we implement the intervention?
- **Cost**: is the intervention affordable for the programme given existing budgetary constraints?

All interventions felt to present any more than minimal risk to participants will be excluded. An explicit trade-off discussion will be held around the maximum financial resources that can be released to fund the testing of one or more intervention (which carries an opportunity cost in terms of the number of people who can be screened) and the minimum ‘meaningful’ improvement that would be required to justify this expenditure. For instance, the local management group may be willing to screen 1% fewer people if attendance rates in the worst-off group improved by >5%. This decision directly informs the next step: agreeing the stopping rule thresholds for the trial (‘x’, ‘y’, and ‘n’ in the three rules below):

1. There is a >**x**% probability that one arm is best.
2. There is a >**y**% probability that the difference between the arms remaining in the trial is <1%.
3. [Optional] A maximum of **n** people have been recruited.

The first two rules will be used for every trial, but the values of x and y will vary depending on the intervention. Some individual trials may introduce a third rule to close trials with indeterminate results after a maximum number of people have been recruited or after a maximum length of time.

The management group will select the thresholds that are most appropriate for the given individual trial, guided by a statistician. The group may accept lower thresholds (and therefore higher risks of type I & II error) for trials of interventions with very low costs, risks, and implementation requirements. For instance, in testing two different versions of a SMS reminder message that are exactly the same cost, the group may use a 51% probability that one arm is best. In contrast, there is a greater imperative to minimise type I and II errors for costly or more risky interventions. The chosen thresholds and the intervention will be reviewed by an independent ethics committee for each individual trial.

We aim to test multiple intervention/service modifications over time in each site e.g. trialling different wording of SMS reminders, or different clinic opening hours, or vouchers of different values – and then take the most effective version to scale across the local programme before repeating the cycle to identify the next intervention/service modifications to test. Individual trials will end once the stopping rules are met. The overall platform trial will close once attendance exceeds 80% for all groups in that particular site.

###### Box 2: Programme management team

The platform trial infrastructure is being set up by the IM-SEEN collaborators, comprised of LSHTM, Peek Vision, COESCA, Kenyan MoH, University of Botswana, NNJS and Dr Shroff’s Charity Eye Hospital using Wellcome Trust and NIHR funds, and in collaboration with national eye care administrators. Decisions around which interventions to test will be made by a multistakeholder group that includes the screening programme funders, implementing partners, and community representatives, with support from LSHTM statisticians. Once the first few interventions have been tested, it is anticipated that the local programme management teams will assume total responsibility for the platform trial process in each country, led by the relevant national decision-makers with responsibility for funding and administering the screening programme in collaboration with local lay representatives and programme implementers. Our ultimate aim is that the broader IM-SEEN process of gathering sociodemographic data, engaging with left-behind groups, identifying interventions, and testing them can be taken to scale across many different sites and services, and that as the approach matures, an increasing number of decisions can be delegated from senior managers to local programme implementers.

#### Types of interventions

This platform trial will be used in each country to test multiple interventions in series i.e. one after the other. It is likely that interventions will be identified that can be administered either at the individual or cluster level. Cluster randomisation will be performed by the teams’ statisticians with pairs of clusters matched by social, geographic, economic and demographic factors. Examples of cluster interventions may include local broadcasts to sensitise populations, new transport services to a given clinic, or changes to the opening times, languages, or locations of clinics.

Examples of individual-level interventions might include vouchers, changes to communication content, wording, timing, and modality (e.g. text message reminder messages), the use of differing visual acuity thresholds, or individual assistance with transport.

We envisage that every consenting person who is referred will be enrolled into the trial that is running at the time they are screened. Our hope is that interventions will lead to a rise in overall attendance in addition to a (larger) rise in attendance among the left-behind population group. This outcome would support the broader goals of proportionate universalism whereby outcomes improve for all, with the greatest gains seen in those with the greatest baseline need.^17^

In some cases, the intervention recommended by the left-behind group and selected for testing may be 100% specific for that group – for instance providing SMS reminders in a new language. In this circumstance, we would not administer the intervention to every person who is referred. Rather, we will restrict that individual trial to the left-behind population group.

Some of these individual-level interventions are digital and could be administered by the Peek software directly after randomisation– for instance by sending different variants of an SMS reminder message, or an electronic voucher via SMS. Other individual-level interventions will require human involvement, such as giving out paper vouchers, or organising transport. Table 2 provides a matrix of example interventions.

Note that this trial will not test any pharmaceutical or medical interventions: the focus is on low-risk service modifications and public health interventions.

This platform trial offers the flexibility of being able to test a number of different interventions under the same master protocol i.e. always using the same population and primary outcomes. Each individual trial that takes place within the overall platform trial will have one or more arms (i.e. different variants/doses of individual interventions) tested against each other and a control. The investigators will not make any efforts to standardise interventions or their delivery as this is a pragmatic trial testing real-world delivery.

The local management group and programme funders will be responsible for obtaining the funding required for each intervention using the resources available to their services. They will be facilitated to apply for external grant funding to cover the costs of interventions where appropriate. We note that many potential interventions such as changing the wording of SMS reminder messages will only incur small marginal costs.

### Discontinuing or modifying interventions

Arms will be discontinued (or modified to remove the risk) if there is evidence that they are harming exposed individuals. We note that only low/negligible-risk service modifications will be tested. Risk will be assessed at the intervention selection stage by a group of researchers, programme managers and lay representatives. Interventions that are deemed to be appropriate will also be independently reviewed by independent ethics committees in each setting before they are implemented in the platform trial.

There are no *a priori* strategies to improve adherence as we are not pre-specifying the interventions.

As our trials will be embedded within routine service delivery, we cannot exclude the possibility that other initiatives will be introduced by local teams before, during, or after individual trials. We will report all programmatic changes that take place during individual trials that could bias our findings.

### Outcomes

This platform trial focuses on testing interventions that improve equitable access to eye services among those identified with a need during screening. We will use attendance as a proxy for access. Our analysis focuses on the population groups found to have the lowest attendance at baseline.

#### Primary outcome

The proportion of people attending triage clinic on their appointed date from the left-behind group, measured using attendance data collected by staff when people check-in.

The left-behind group will be identified at baseline as part of the ‘identify’ stage of the IM-SEEN process. This group will be constituted of the group(s) with the lowest baseline attendance rates that collectively constitute at least 10% of the total population. A focus on left-behind groups is important to programme managers who are trying to close gaps, extend health service coverage, and ensure that their services do not exacerbate existing inequalities.

When referred participants check-in at ophthalmic clinics, their attendance status is recorded by administrative staff using the Peek app, which automatically updates a central database that holds records of each participant’s eye care need, sociodemographic characteristics, arm allocation, and attendance status at the ophthalmic clinic on the appointed date. Our Bayesian algorithm will review the attendance data for every referred participant every 72 hours and calculate the probability of attendance within each arm. In our modelling we have estimated that 100 people will be referred every 72 hours. This aligns with what we have observed in India and Kenya where approximately 1,000 people are screened per day, of whom approximately 1/3 are referred. We have stipulated that the left-behind group will include at least 10% of the total population (i.e. 100 people every 72 hours). In programmes where fewer people are referred every 72 hours, the interim analysis window will be extended as appropriate.

#### Secondary outcome

The proportion of people attending triage clinic on their appointed date across the entire population, measured using attendance data collected by staff when people check-in.

If an intervention is found to increase attendance among the left-behind group, we also want to check whether there has been an impact on the overall mean attendance rate. This is to hedge against adopting an intervention that improves access for the left-behind group but leads to a large overall fall in attendance across the entire programme. We will use absolute percentage differences in attendance for comparisons between the left-behind and general populations exposed to the intervention.

### Participant timeline

This platform trial is embedded within routine screening programmes. From the individual participant perspective, they will flow through the screening programmes as normal; participants will present and have their eyes checked by a first-line screener either in their own home, at a school, local clinic, or community meeting place, depending on the setting. The screener will ask a series of sociodemographic questions and perform a ‘tumbling E’ visual acuity assessment, all using the Peek smartphone app. Those who screen positive will be referred to a local triage centre where their eyes will be re-checked by a more highly skilled practitioner and treatment will be delivered. Those requiring more advanced care will be referred on to the appropriate service provider.

Some programmes use a roaming team of screeners who visit communities sequentially. Others train screeners who remain in one location. Table 3 summarises the two approaches.

**Table 3:** Different screening programme approaches.

|  | **Outreach screening model** | **Primary care screening model** |
| --- | --- | --- |
| **1. Community-based screening** | Outreach screeners trained to use the Peek app attend schools/local venues and screen those who are present before moving to the next location | Primary care staff are trained to use the Peek app to opportunistically screen and refer those who present to primary care |
| **2. Referral to triage** | Those who screen negative (i.e. with no eye health need) are discharged. Those who screen positive provide their contact details and answer a series of socioeconomic questions. They are then referred to triage. |  |
| **3. Triage, basic treatment, & further referral** | An ophthalmic nurse checks-in attenders using the Peek app and then performs an eye examination. Simple treatments are provided for basic issues (e.g. eye drops for conjunctivitis). Other cases are referred for refraction and/or further care. Referral status is recorded in the Peek app. |  |
| **4. Specialist treatment** | A receptionist checks-in those who present to receive refraction or further ophthalmic treatment using the Peek app at the secondary or tertiary clinic. |  |

In some settings, triage will be co-located with screening. In others it will be co-located with the provision of refractive services, and in others it will be co-located with refraction and all other specialist treatment providers i.e. in a hospital. Figure 4 shows the three different configurations of screening, triage, and treatment.

**Figure 4:**
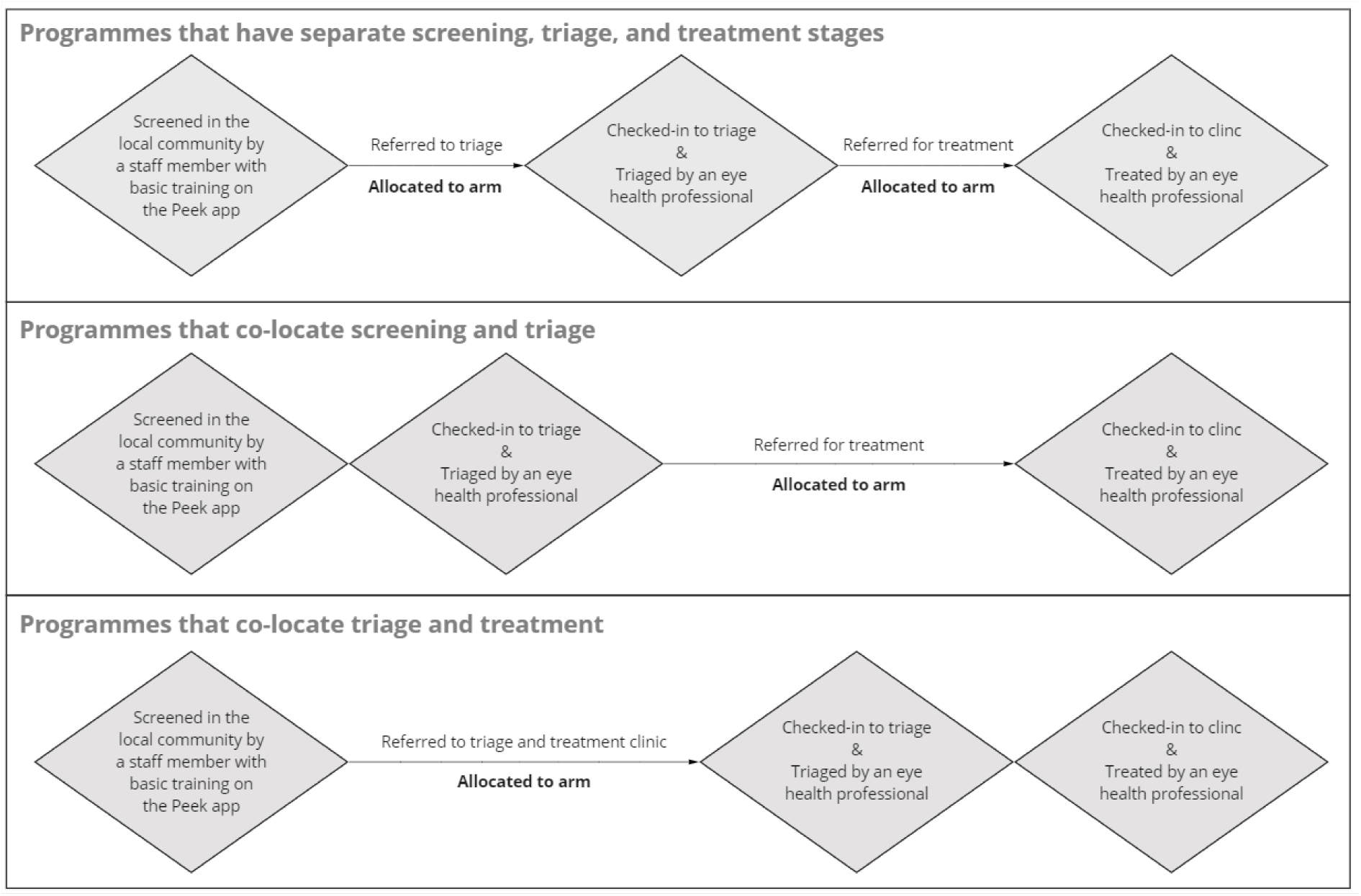
Flow through a generic screening programme for those requiring treatment

Most programmes aim to progress to a model of co-locating triage with screening or treatment in order to reduce the appointment burden on participants and minimise loss-to-follow-up. In the former case, participants testing positive at screening are ‘referred’ to a room next door for triage. In the latter case, they are given an appointment to attend a central triage & treatment clinic, commonly 1-2 weeks after screening. In most programmes SMS reminders are sent on the date of referral and the day before the appointment. Interventions will be allocated by the algorithm at the point of referral.

### Sample size

As we are using stopping rules, will not pre-specify a minimum sample size or estimate effect sizes for the intervention arms. Instead, participants will be continually recruited until we reach a pre-determined maximum sample size or sufficient data accrue to trigger one or more of the other stopping rules. Triallists have argued that this approach is more “efficient, informative and ethical” than traditional fixed-design trials as this approach optimises the use of resources and can minimise the number of participants allocated to ineffective or less effective arms.^18^ Every 72 hours the algorithm will review the attendance data and calculate the probability of attendance within each arm.

#### Operating characteristics for individual trials of interventions administered to individual participants

Based on extensive scenario modelling, we have decided to use the following stopping rules for individual trials that test interventions administered to individuals (rather than clusters):

1. There is a >x% probability that one arm is best i.e. there is a >0% difference between the arms. (Default x = 95%)
2. There is a >y% probability that the difference between the best arm and the arms remaining in the trial is <1%. (Default y = 95%)

Individual trials may include a third ‘ceiling’ stopping rule around a maximum length of time that the trial will run for, or a maximum sample size, depending on the context. For instance, there may only be funding to run a particular service modification for 12 months, or there may only be capacity to trial a new intervention for the first 10,000 people. The default ceiling will be 10,000 participants.

Each individual trial will end once one or more of the stopping rules has been met. At that point the superior arm will become the new standard care in the programmes(s) where it was tested. The overall platform trial will be stopped once attendance reaches or exceeds 80% for every sociodemographic group in a given site. Figure 5 provides a visual representation of how the trial will run, including the point at which new interventions can be added.

**Figure 5:**
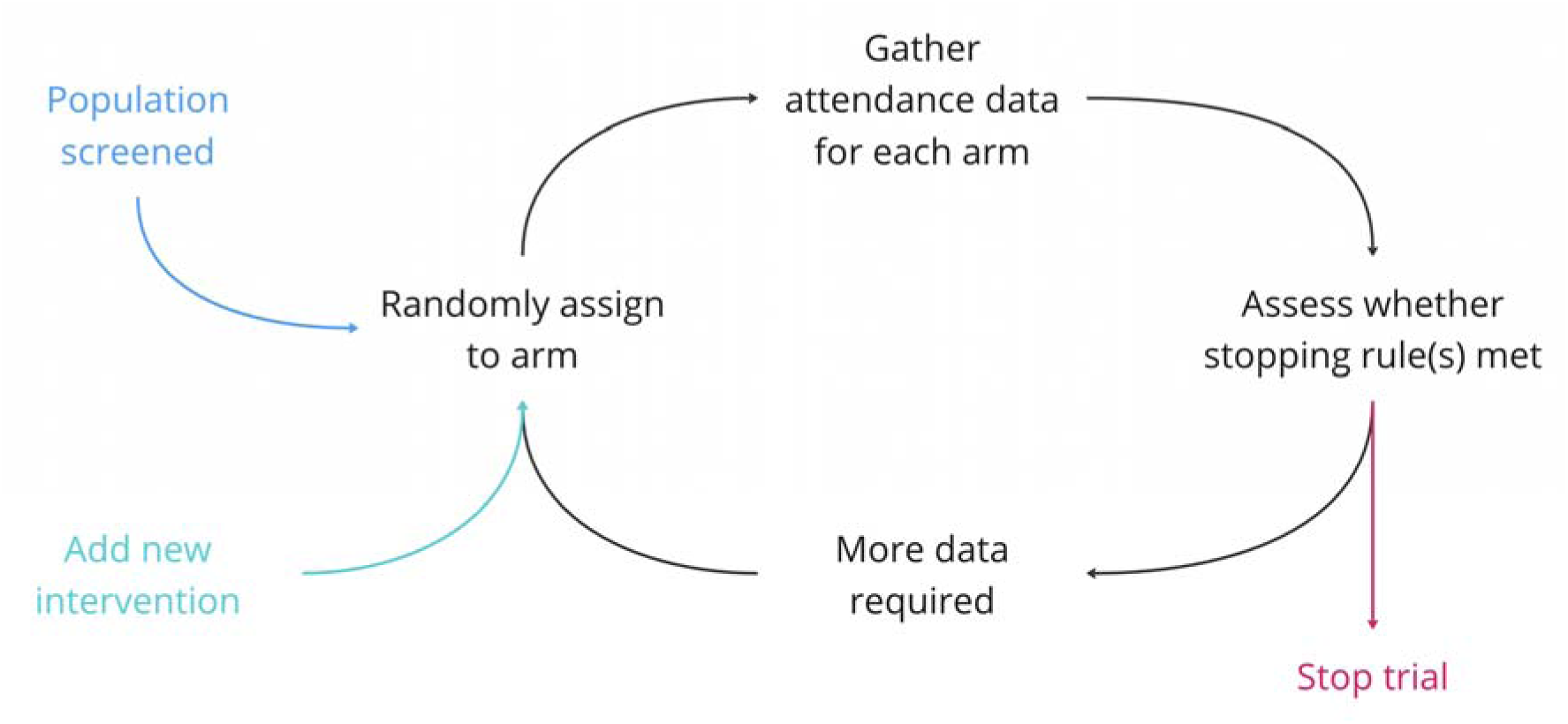
Platform trial schematic

We conducted simulations to estimate the impact of the early stopping rules on error rates and sample sizes. For both rules, 95% threshold values were used as default. We assumed a fixed 1:1 ratio for two-arm trials where the control arm had 50% outcome rate and the intervention arm has an effect difference of d, ranging from 0% to 5%. A total of 1,000 simulations were conducted for each value of d, and we assumed that interim analysis would take place for every 100 outcomes observed.

In this trial, we prioritize high statistical power (1-β). Minimizing β will protect against the risk of incorrectly identifying an inferior arm as a winning arm. Simulation results show that the expected power in our trial will be at least 98% when an intervention arm is more effective than the control arm by a difference of 3% or greater. When the winning arm is only marginally more effective by a difference of 1%, our trial will still ensure a statistical power of 92%, which is greater than the power of 80% used in most conventional trials. It is noted that the high statistical power in our trial comes at the cost of increased chance of committing type I error. Furthermore, it will take longer to run the trial to find smaller differences. When there is no difference between arms, we expect 32% chance of making false positive conclusions (Table 4). But we will treat the risk of committing type I error as not a major concern because we expect no or minimal harm in selecting either of the two arms with equal effectiveness.

**Table 4.** Expected error rates and sample size, by true effect difference between arms (d)

| True effect difference between arms (d) | Type I error ($\alpha$) | Type II error ($\beta$) | Median sample size [IQR] |
| --- | --- | --- | --- |
| 0% | 32.1% |  | 19,950 [3075,43525] |
| 1% |  | 8.4% | 8,150 [2500,22650] |
| 2% |  | 3.1% | 3,800 [1500,8100] |
| 3% |  | 1.8% | 2,100 [1000,4100] |
| 4% |  | 0.4% | 1,600 [900,2700] |
| 5% |  | 0.6% | 1,200 [700,2000] |
| 10% |  | 0% | 500 [400,800] |
| 15% |  | 0% | 300 [300,400] |
| 20% |  | 0% | 200 [200,300] |
| 25% |  | 0% | 200 [200,200] |

The values of posterior probabilities specified in rules 1 and 2 will be determined by our research team at the start of each individual trial. The default values will be 95% as above, however it might be appropriate to use lower thresholds for interventions where the costs and risks are negligible, and higher thresholds when the costs and/or risks are high. For example, to decrease the chance of committing type I errors, the probability threshold in rule 1 will be increased from 95% to a higher value (Figure 6).

**Figure 6.**
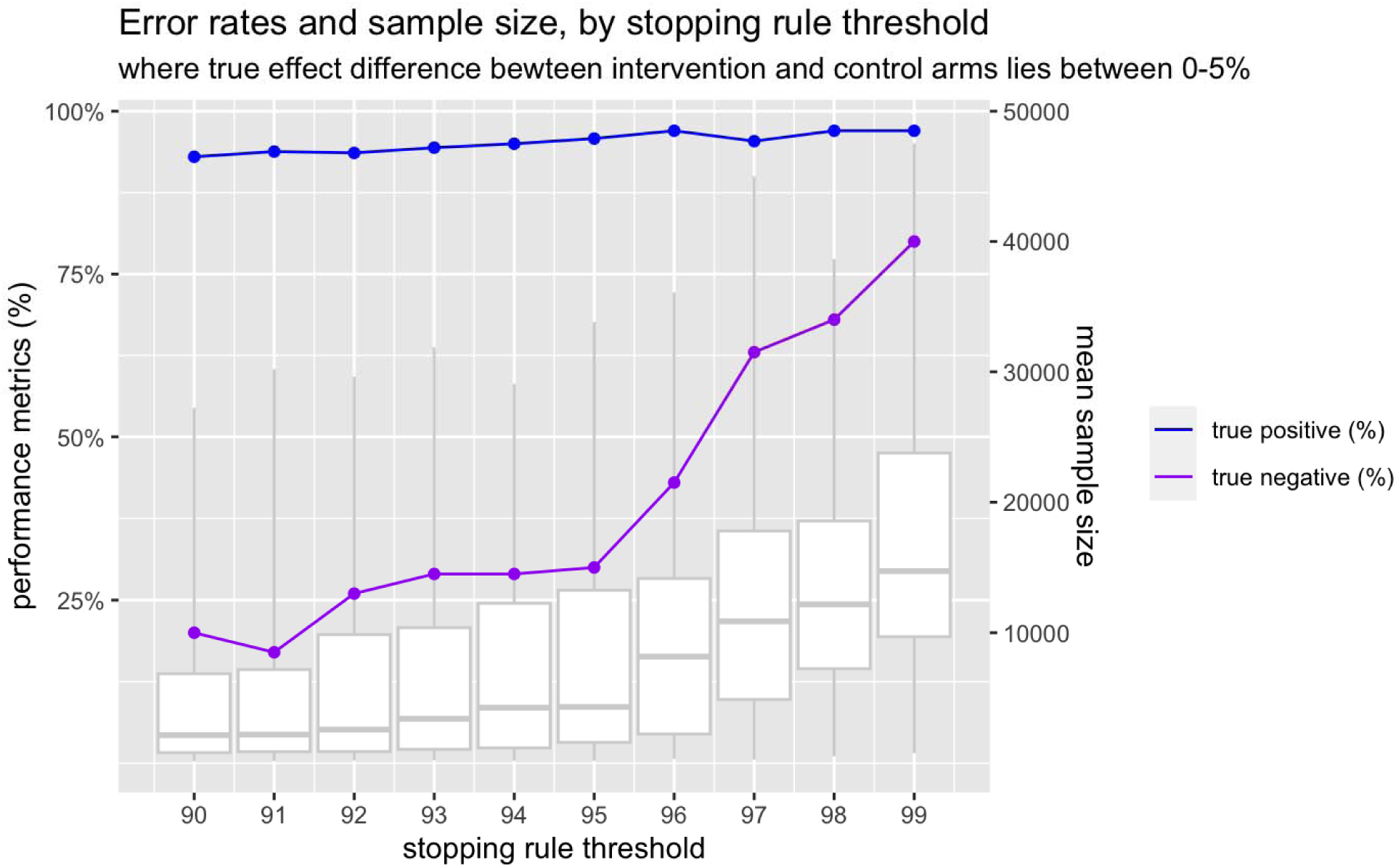
Expected error rates and sample size, by changing stopping rule threshold

#### Interventions administered to clusters

Where the chosen intervention can only be implemented in clusters rather than randomising individuals to receive the intervention, the local management team will be convened to develop a design tailored to the intervention. An important factor to account for in any design will be determining how much the outcome varies by cluster and how large each cluster is. For cluster-level interventions it is likely we will carry out a more traditional approach with a fixed number of clusters randomised before declaring one arm the winner. The number of clusters randomised will be based on the intra-cluster correlation, the current attendance rate, the size of the clusters and the effect size for which we want to be powered to detect.

### Recruitment

As the trial is pragmatic, the responsibility for recruiting screening participants lies exclusively with local programme managers. Programme implementers will enrol participants by seeking consent from all those who require referral for further assessment and care.

### Allocation

#### Sequence generation

We will use computer-generated random numbers to generate the allocation sequence and assign all consented, referred participants to intervention arms, with equal numbers of participants in each arm. Where appropriate blocking will be used with blocks between 4-12. Stratification will be used where appropriate.

#### Allocation concealment mechanism

For interventions delivered to individuals, the allocation sequence will be generated within the Peek system in real-time, as participants are referred. As human trial managers are not involved in allocation there is no need for concealment.

For cluster trials these will be done randomly. Restricted randomisation will likely be used in this scenario to achieve balance between arms.

### Implementation

The algorithm will be set up so that it can implement digital interventions such as SMS messages without human investigators being exposed to the allocation status of individual participants. For interventions that require human intervention – such as providing transport, chaperones, or physical vouchers, implementers will be informed of individual participants’ assignment status via the Peek app at the stage that intervention needs to be delivered.

### External independent review of interventions prior to implementation

As and when new interventions are selected for testing, they will need to be externally reviewed by an independent national ethics committee to ensure that the intervention(s) do not pose undue risk. The platform trial is designed to test low/negligible risk service modifications. Coupled with the fact that the master protocol will already have receive ethical approval, this should enable rapid/expedited ethical review of new interventions rather than full committee review. Table 5 summarises example interventions and risk thresholds.

**Table 5:**
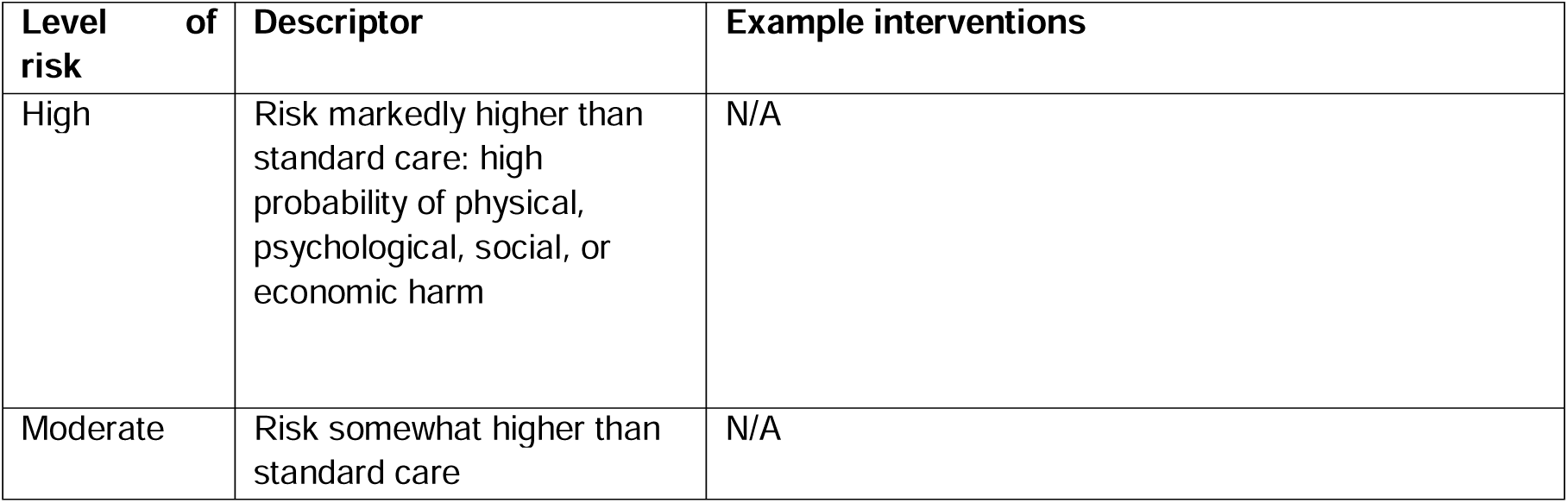
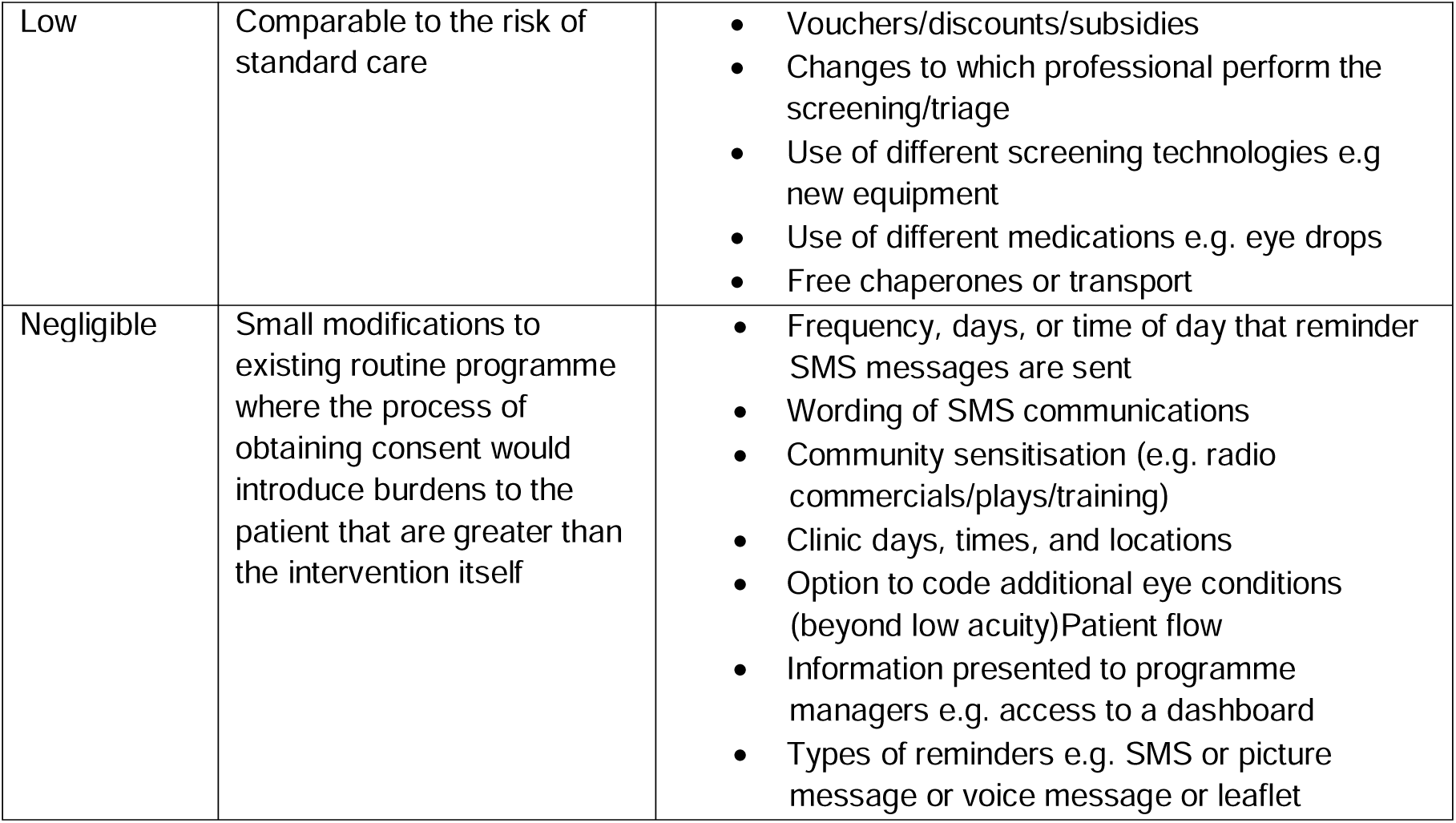
Risk thresholds and example interventions.

| **Level of risk** | **Descriptor** | **Example interventions** |
| --- | --- | --- |
| High | Risk markedly higher than standard care: high probability of physical, psychological, social, or economic harm | N/A |
| Moderate | Risk somewhat higher than standard care | N/A |
| Low | Comparable to the risk of standard care | • Vouchers/discounts/subsidies<br>• Changes to which professional perform the screening/triage<br>• Use of different screening technologies e.g. new equipment<br>• Use of different medications e.g. eye drops<br>• Free chaperones or transport |
| Negligible | Small modifications to existing routine programme where the process of obtaining consent would introduce burdens to the patient that are greater than the intervention itself | • Frequency, days, or time of day that reminder SMS messages are sent<br>• Wording of SMS communications<br>• Community sensitisation (e.g. radio commercials/plays/training)<br>• Clinic days, times, and locations<br>• Option to code additional eye conditions (beyond low acuity) Patient flow<br>• Information presented to programme managers e.g. access to a dashboard<br>• Types of reminders e.g. SMS or picture message or voice message or leaflet |

Once the master adaptive platform trial has received ethics approval, individual interventions will be submitted as amendments to the master APT in the form of new appendices. Individual trials will not commence until ethics approval has been received from LSHTM and the relevant local ethics committee(s).

### Masking

#### Who and how

Once assigned by the algorithm, each participant’s online record will automatically update to display which arm they have been allocated to. Participants will not be masked to assignment. For interventions that require human delivery (e.g. handing out a paper voucher), implementers will be able to view allocation status out of necessity. Outcome assessment will be performed by a different group - those responsible for checking-in participants at triage clinic. No steps will be taken to mask these staff to participant allocation status. Ongoing interim data analysis will be performed by the Bayesian algorithm every 72 hours.

#### Unmasking

Human investigators and programme managers will not be able to access data on allocation of participants to specific arms unless they are involved in delivering an intervention.

The Data Safety and Monitoring Committee (DSMB) will have access to all data at any point and for any reason, including to unmask assignment if required. The trial steering committee members will only be able to access these data as per the adverse event protocol outlined below.

### Data Collection

#### Data collection methods

As stated above, outcome assessment (attendance at clinic) will be recorded when participants check-in at clinic on their appointed date. Each participant’s attendance status will be recorded on their central record.

#### Retention

There are no plans to promote participant retention and complete follow-up.

#### Data management

All data entry will be performed by programme staff as part of routine screening and clinical care. See the data management plan for further information about coding, security, and storage.

### Statistical methods

All analysis will be conducted using R. Baseline characteristics of all participants will be described as mean (SD) or median (IQR) for categorical variables, or as frequencies and proportions for continuous variables.

During this adaptive trial, clinic attendance in each arm will be assessed using Bayesian methods. At each prespecified interim analysis point, a binomial distribution of outcome will be described for each arm using the total number of participants allocated to the arm and the number that attended at clinic. The binomial distribution will be combined with a prior distribution to update the posterior distribution of each arm. A regularizing prior of beta(100,100) will be applied to reduce overfitting until a reliable amount of data is accrued. A Monte-Carlo simulation will be used to update posterior distributions at each interim analysis point. Posterior probabilities will be calculated and compared to the stopping rules as to whether the trial should continue into the next day or end early. If there is sufficient evidence to meet one of the stopping rules, the trial will terminate and proceed to the final analysis stage.

Upon completion of the trial, a complete case analysis will be performed on all eligible participants in the trial on an intention-to-treat basis. The primary endpoint of the trial is clinic attendance the left-behind subgroups after randomization. Within a selected subgroup, the primary analysis will use beta-binomial models to estimate the posterior distribution of attendance in each arm. Posterior probabilities will be calculated to compare the proportion of attendance between arms and to identify an arm that results in the highest likelihood of attendance. For the secondary endpoint, beta-binomial models will also be used but expanded to all participants in the trial. A more detailed description of the statistical methods will be reported as open access as a separate statistical analysis plan.

### Equity analyses

The primary aim of the platform trial is improving equity. We focus on attendance rates in the left-behind group, and also look at how attendance rates in this group compare to those among the entire population.

### Non-adherence and missing data

Missing data is not a problem because the outcome is attendance. Non-adherence will depend on the intervention. We will use intention-to-treat analysis.

An independent Data and Safety Monitoring Board (DSMB) will be appointed in each country with the primary aim of assuring safety of participants in the trial(s). The DSMBs will advise the steering committee and sponsor on continuation or stopping of the trial(s) based on safety and efficacy considerations. Each DSMB will have three members, all independent of the running of the trial, and all with relevant clinical and epidemiological experience. Each DSMB will operate independently of the study sponsor and the steering committee. Each DSMB will confirm their own specific meeting arrangements and draw up their own charter, working from the template produced by the Damocles Study Group.^19^ It is proposed that each DSMB would meet prior to the beginning of each individual trial conducted under the platform protocol, one third of the way through, and at the end of each individual trial, to assess the safety of the trial procedures. Each DSMB will agree the way it will monitor the data, what it requires from the investigators in this respect and will communicate this to the PIs. All data can be interrogated remotely in real-time. The DSMB may visit the study coordination centre to assess data management, record keeping and other important activities. Each DSMB will determine the manner in which it will monitor the data, what it requires from the investigators in this respect and will communicate this to the PIs.

### Botswana DSMB

- Billy Tsima
- Lemphi Moremi
- Mantate Manyothwane

### Kenya DSMB

- Nyawira Mwangi
- D Stephen Gichuhi
- Moses Mwangi

### Nepal

- Sabina Shrestha
- Sanjib Mishra
- Rajiv Ranjan Karn

### India

- Shalinder Sabherwal
- Javed Nayab
- Atanu Majmudar

### Consent

Written informed consent will be sought by screeners during screening - at the point that participants are identified as having an eye care need and referred on for further care. Consent will be recorded either on paper forms or by using an electronic tick box (as appropriate for low-risk trials). Whichever format is used, consent status will be recorded on the Peek app.

Participants will be given the contact details of the research managers and will be free to leave the trial at any time. There will be no remuneration for participants.

### Patient and public involvement

Lay people and community advisory committees have reviewed and contributed to the development of this protocol. The interventions that the platform trial will test will be derived from engagement with affected groups. Lay representatives will assist with interpretation and publication of the trial findings.

### Adverse event reporting and harms

An adverse event (AE) is defined as any untoward medical occurrence in a patient or study participant. All adverse events will be reported. Depending on the nature of the event the reporting procedures below will be followed. Any questions concerning adverse event reporting will be directed to the study coordination centre in the first instance. The flow chart below has been provided to aid the reporting of adverse events.

#### Non-serious AEs

All non-serious AEs will be reported to the study coordination centre and recorded in a dedicated AE log within 72 hours. The entry must state the patient ID, date and time of AE, nature, and relation to the intervention, if any. The AE should also be reported to the data and safety monitoring committee within 72 hours. AE logs will be stored on a secure, password-protected file on a LSHTM computer.

#### Serious AEs

Serious Adverse Events (SAEs) will be reported to the PI and study coordination centre within 24 hours of the local site being made aware of the event (Figure 5). The PI will report the event to the data safety monitoring committee within 48 hours and include it in the study safety report.

**Figure 5:**
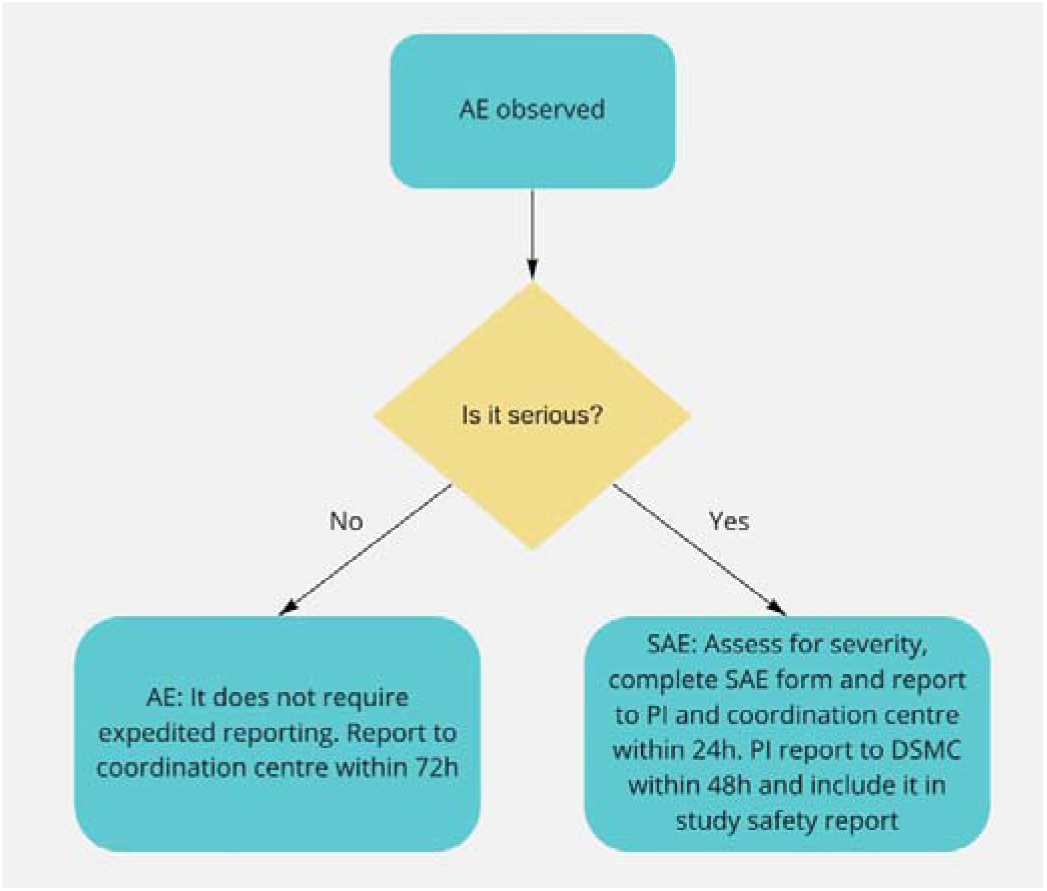
Approach for managing adverse events

An SAE form will be completed and submitted to the PA and study coordination centre with details of the nature of event, date of onset, severity, corrective therapies given, outcome and causality. All SAEs whether expected, suspected or unexpected will be reported to regulatory bodies and the trial DSMB within 48 hours of occurrence. The responsible investigator will assign the causality of the event. All investigators will be informed of all SAEs occurring throughout the study. If awaiting further details, a follow up SAE report should be submitted promptly upon receipt of any outstanding information.

Any events relating to a pre-existing condition or any planned hospitalisations for elective treatment of a pre-existing condition will not need to be reported as SAEs.

### Limitations

We have chosen to use a prioritarian approach that focuses on left-behind population groups. This prevents a situation where we accept an intervention that improves mean attendance but is associated with a decline among left-behind groups. However, this approach does not hedge against the slope of inequality worsening. Unfortunately, using a proportionate approach where we assess whether gains in each group are proportionate to their initial need would risk attributing success to our intervention rather than the more likely detection of regression toward the mean.

Our estimate of the probability/proportion will be biased because, on average, the stopping rules will be triggered at a ‘local peak’. As such, we will be able to identify that, say, A is better than B, but the estimate of the attendance rate in A will be an overestimate.

We use attendance as a proxy for access. Whilst this is the closest hard indicator available, the semantic implication of the term places responsibility on people rather than clinical systems or societal structures. We will counterbalance this in the language that we use to talk about barriers and in the framing of interventions in our individual study writeups. We also note that we focus on a proximal indicator that does not always correlate well with receipt of high-quality care, or good clinical outcomes. We decided to focus on access for three main reasons; first it aligns with the conceptual narrative of Universal Health Coverage and ‘leaving no one behind’, second attendance data are already routinely collected and available for every single person who is referred, and third, internal Peek data suggests that the ‘fall off’ gap between those who are referred but do not attend is much larger than other gaps e.g. the proportion of those who attend but do not receive appropriate care, or the proportion of those who receive appropriate care but do not experience improved health outcomes.

## Supporting information

CONSORT Statement

## Data Availability

All data produced in the present study are available upon reasonable request to the authors.

## Dissemination

Each individual trial will have its own protocol that will be published online. The results of each trial will be immediately fed back to the relevant programme managers. Findings will also be shared with wider stakeholders, including eye care professionals, policymakers, and community representatives at dedicated dissemination meetings. We will write-up all individual trials for publication in the peer-reviewed literature and share lay-friendly summaries via social media.

## Appendix: CONSORT checklists

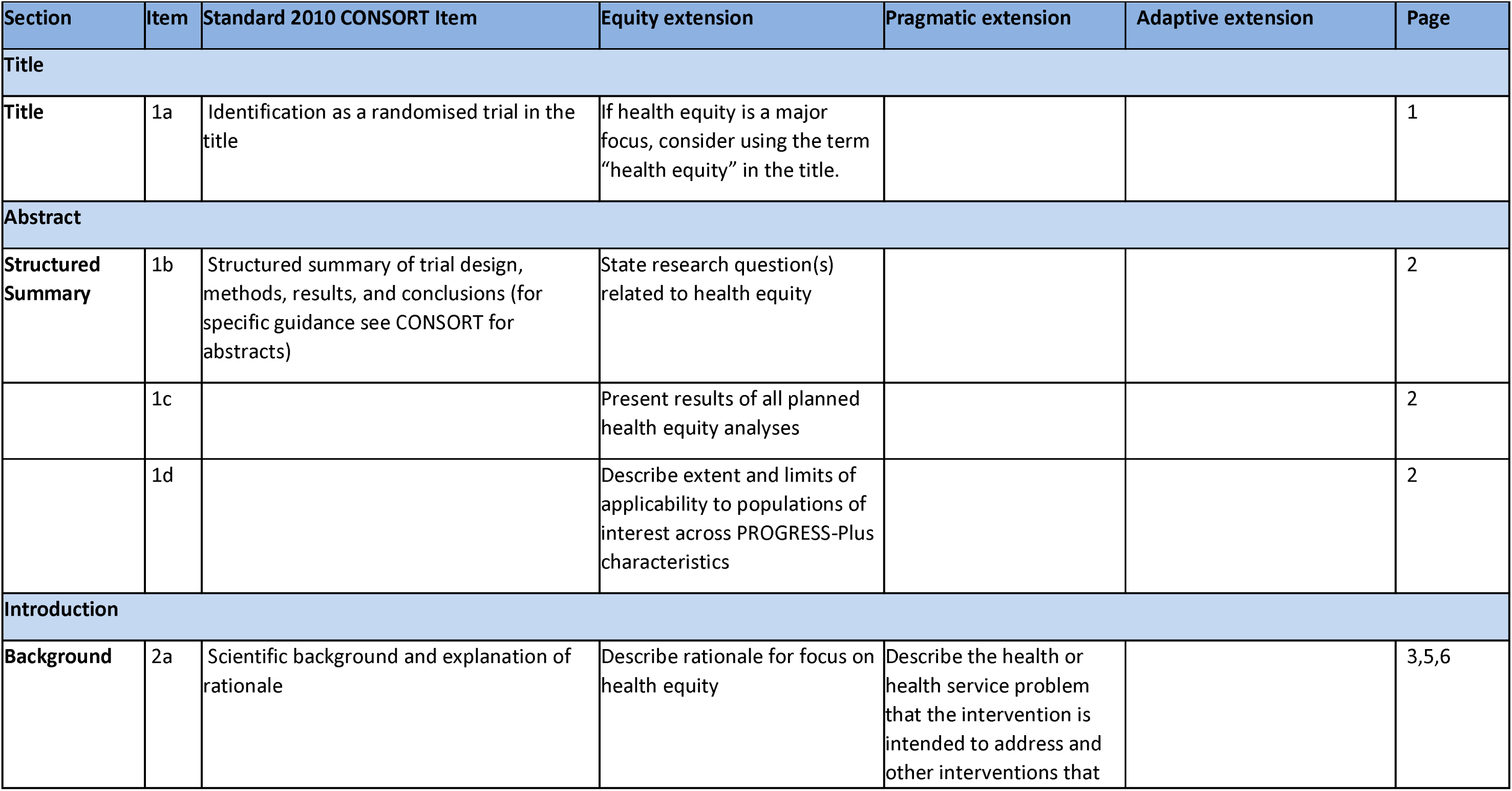
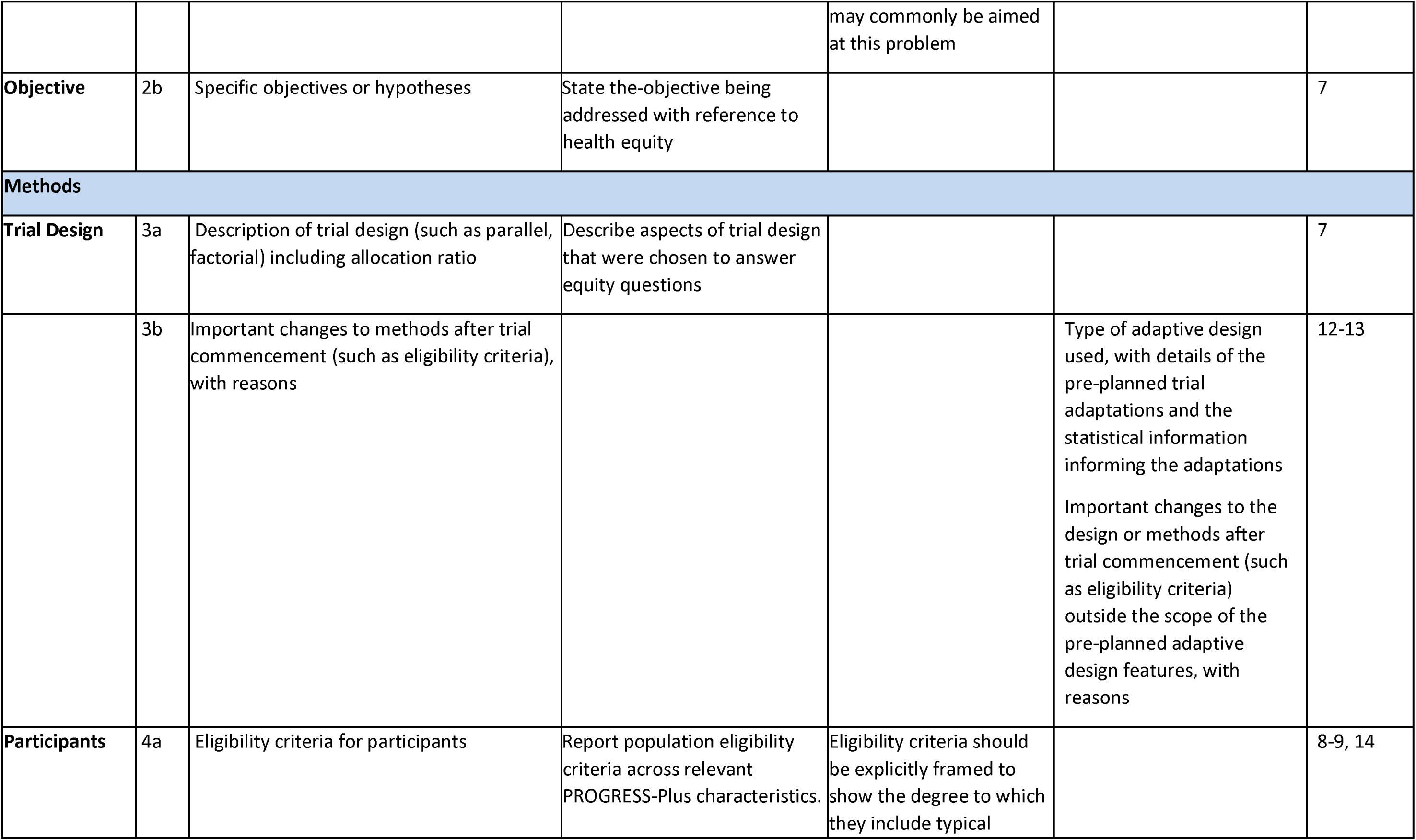
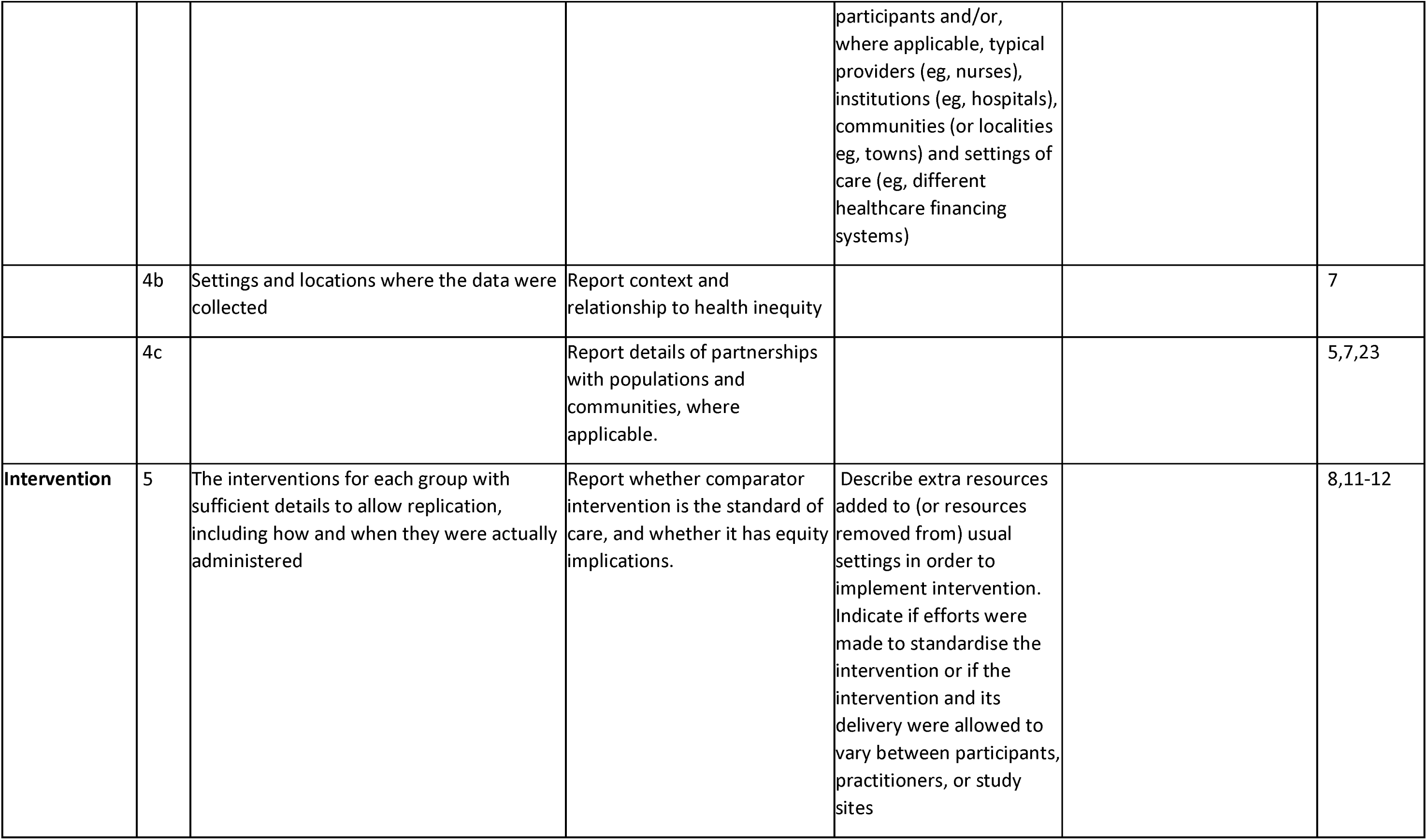
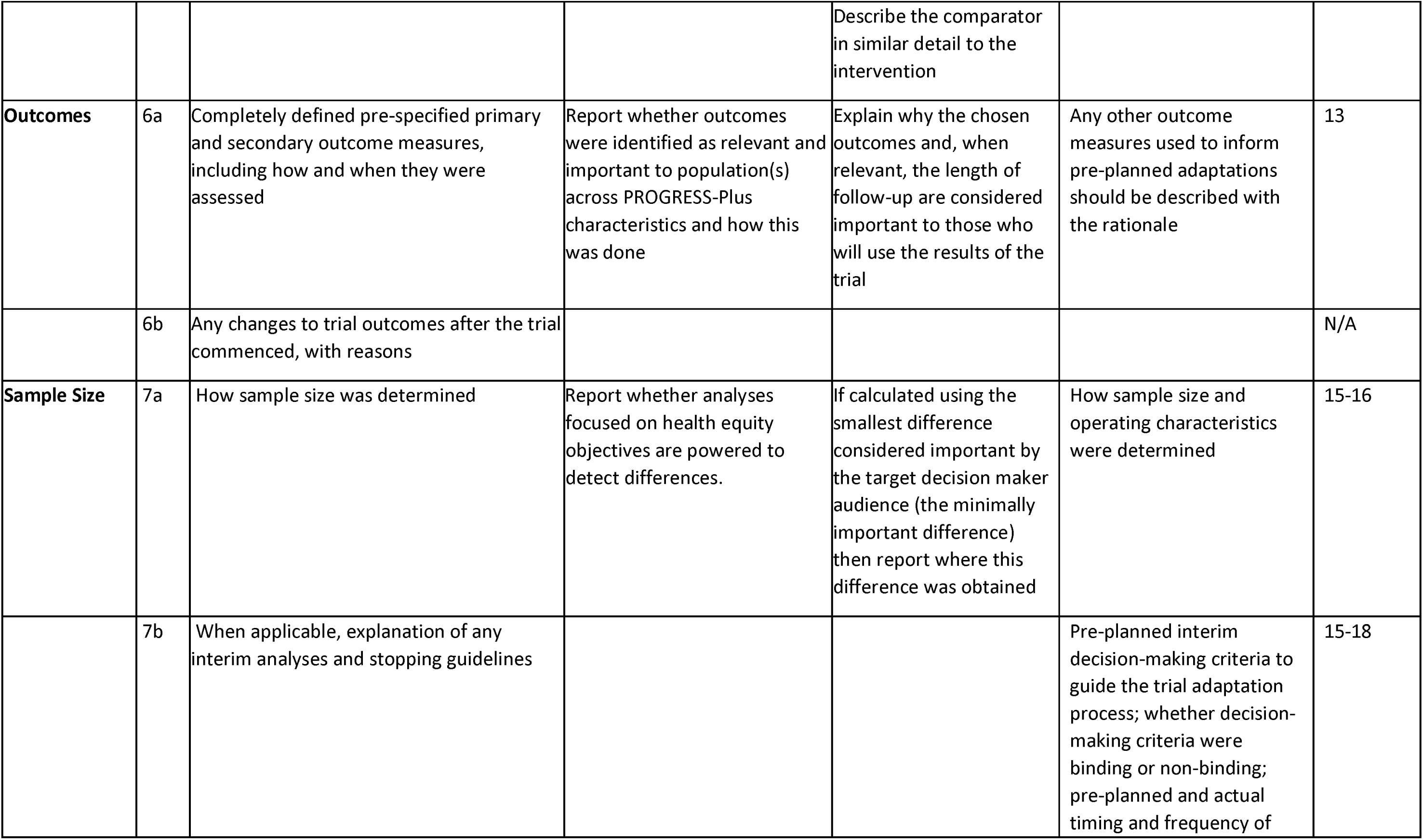
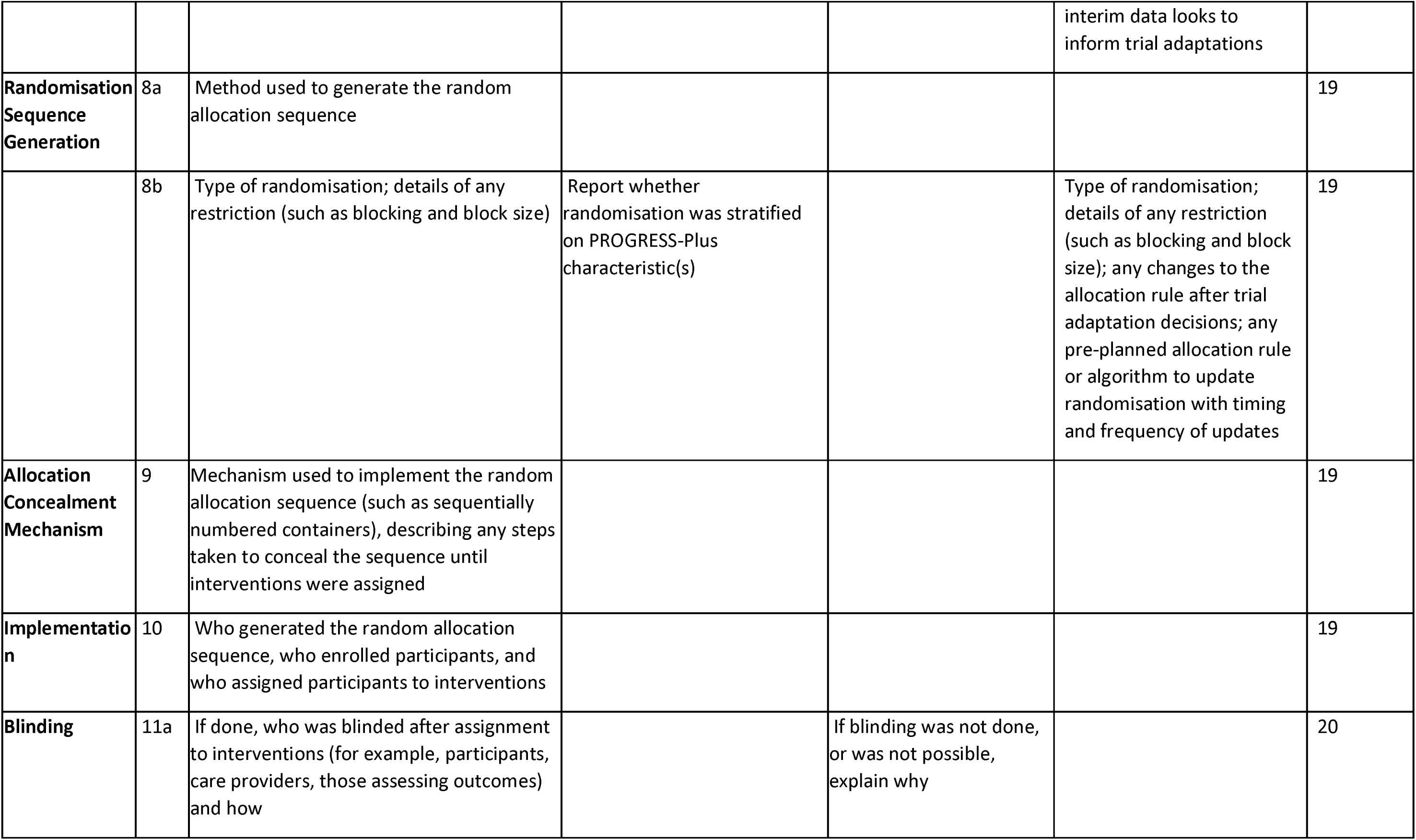
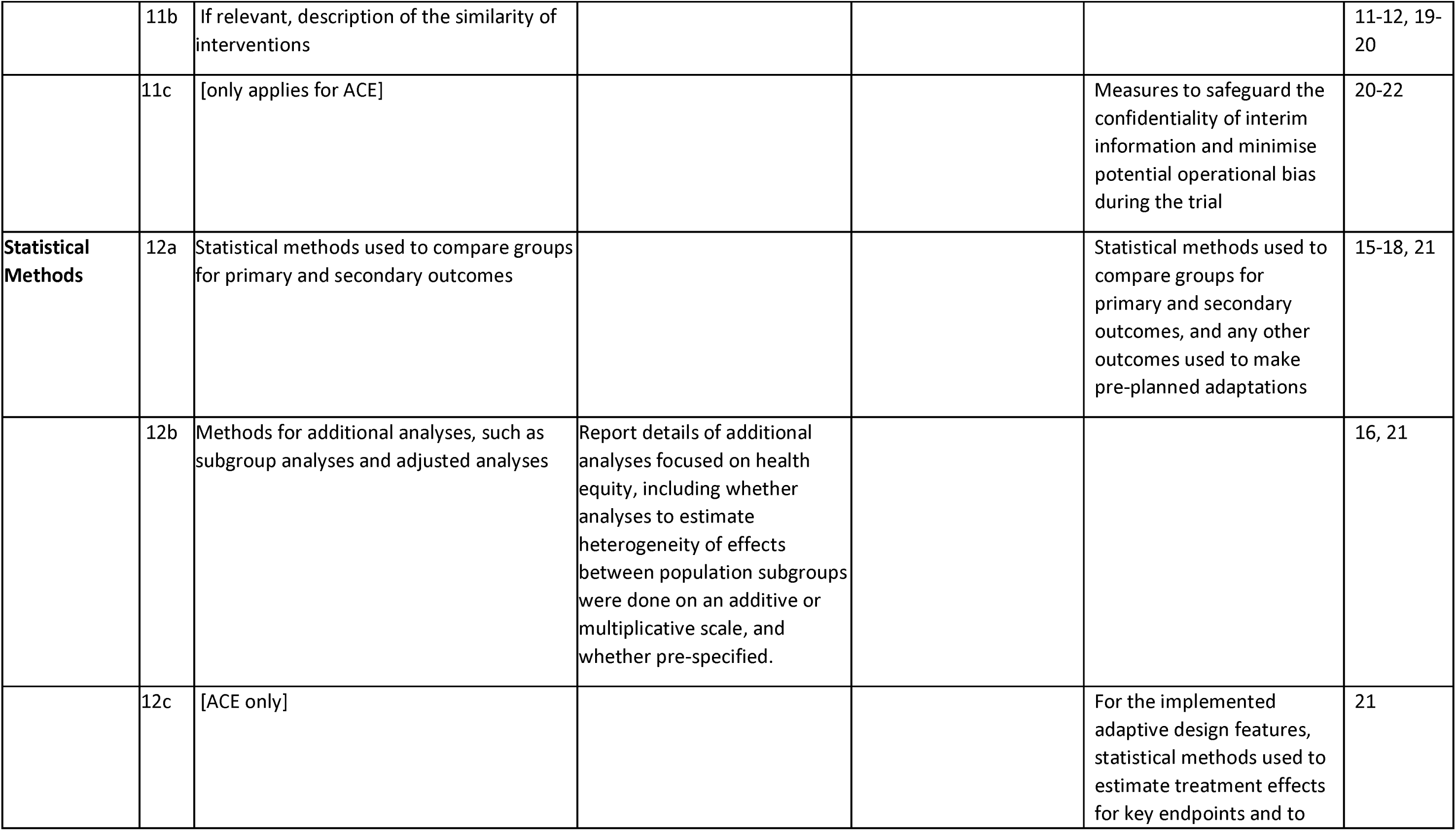
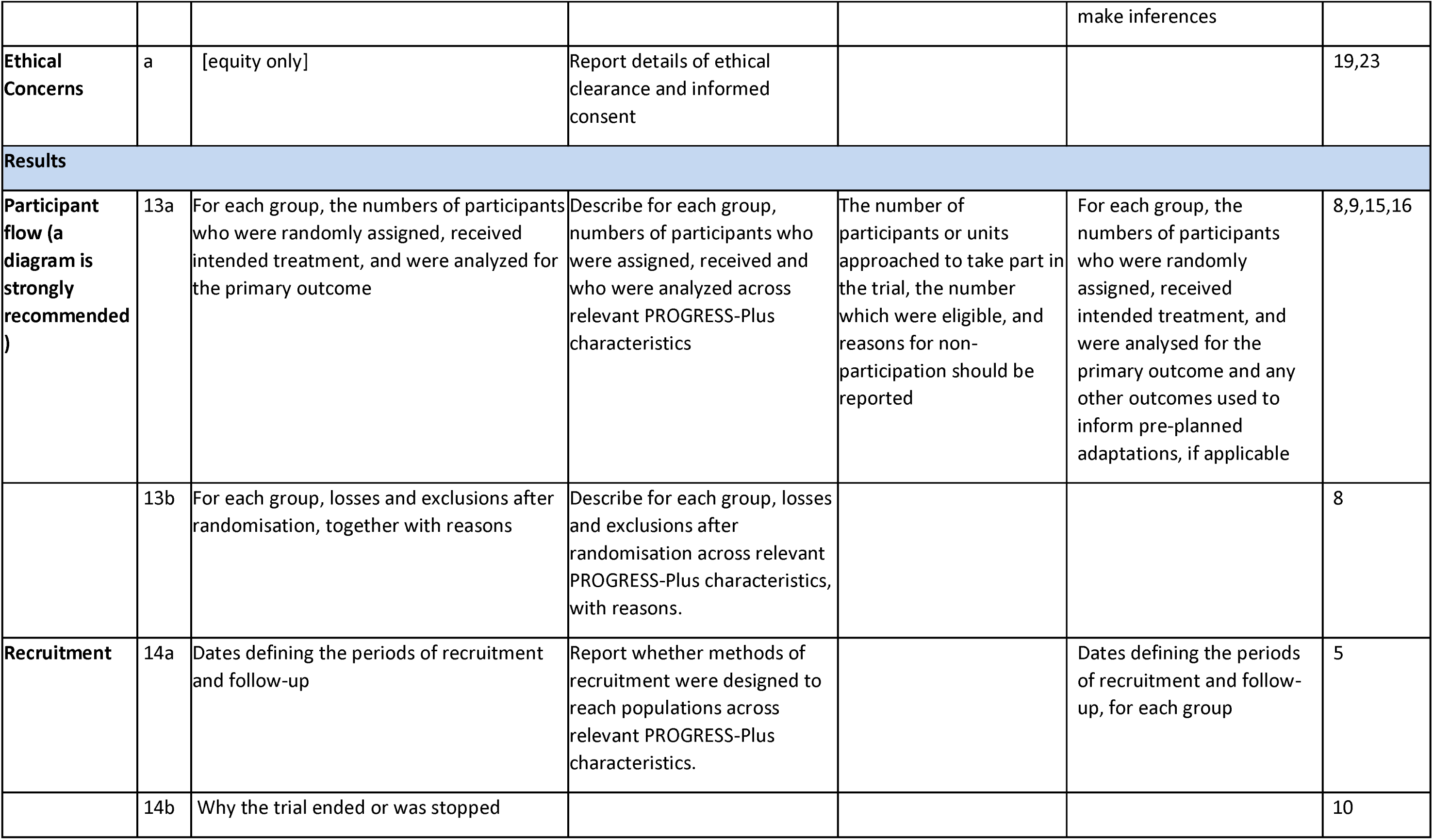
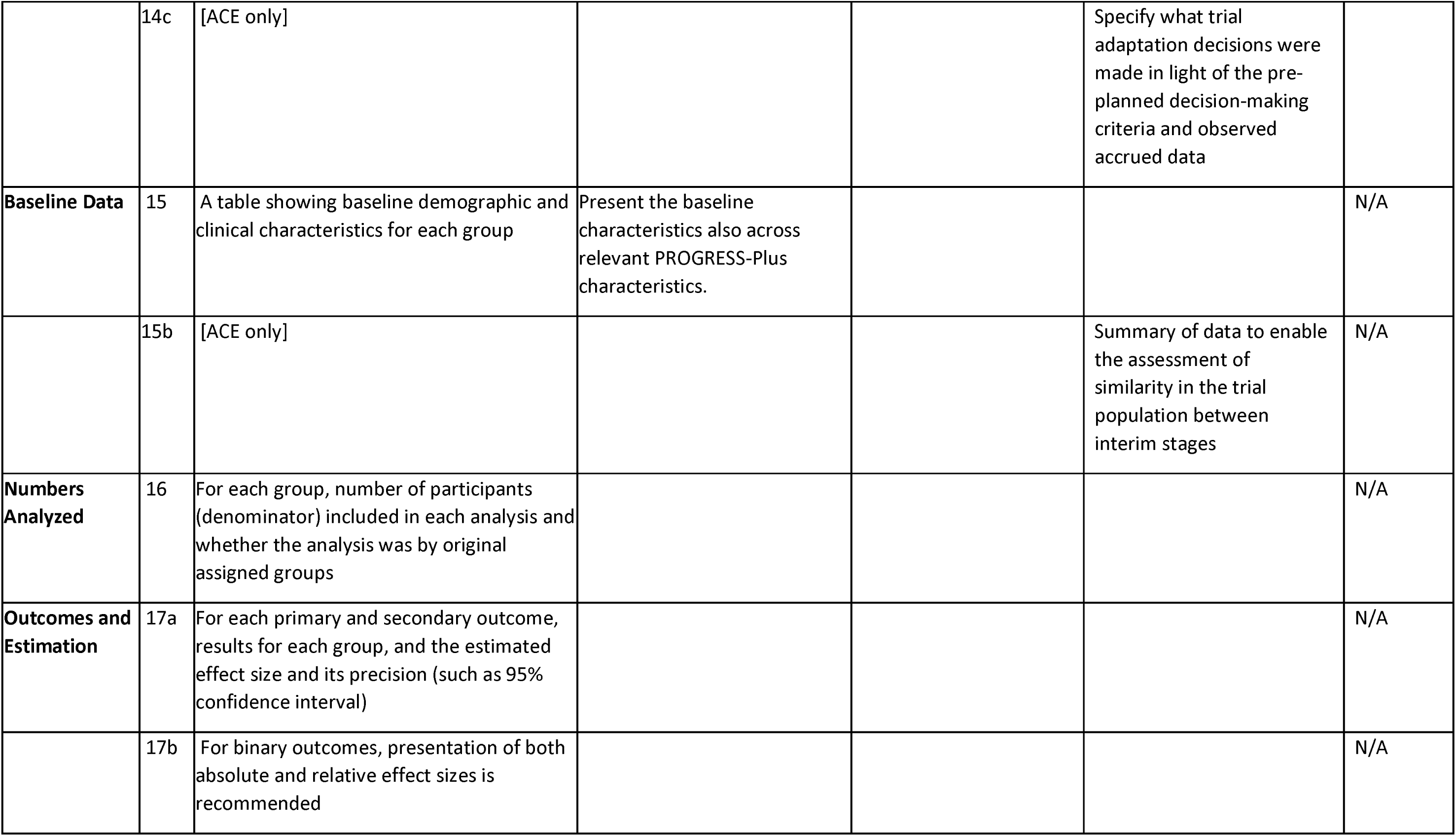
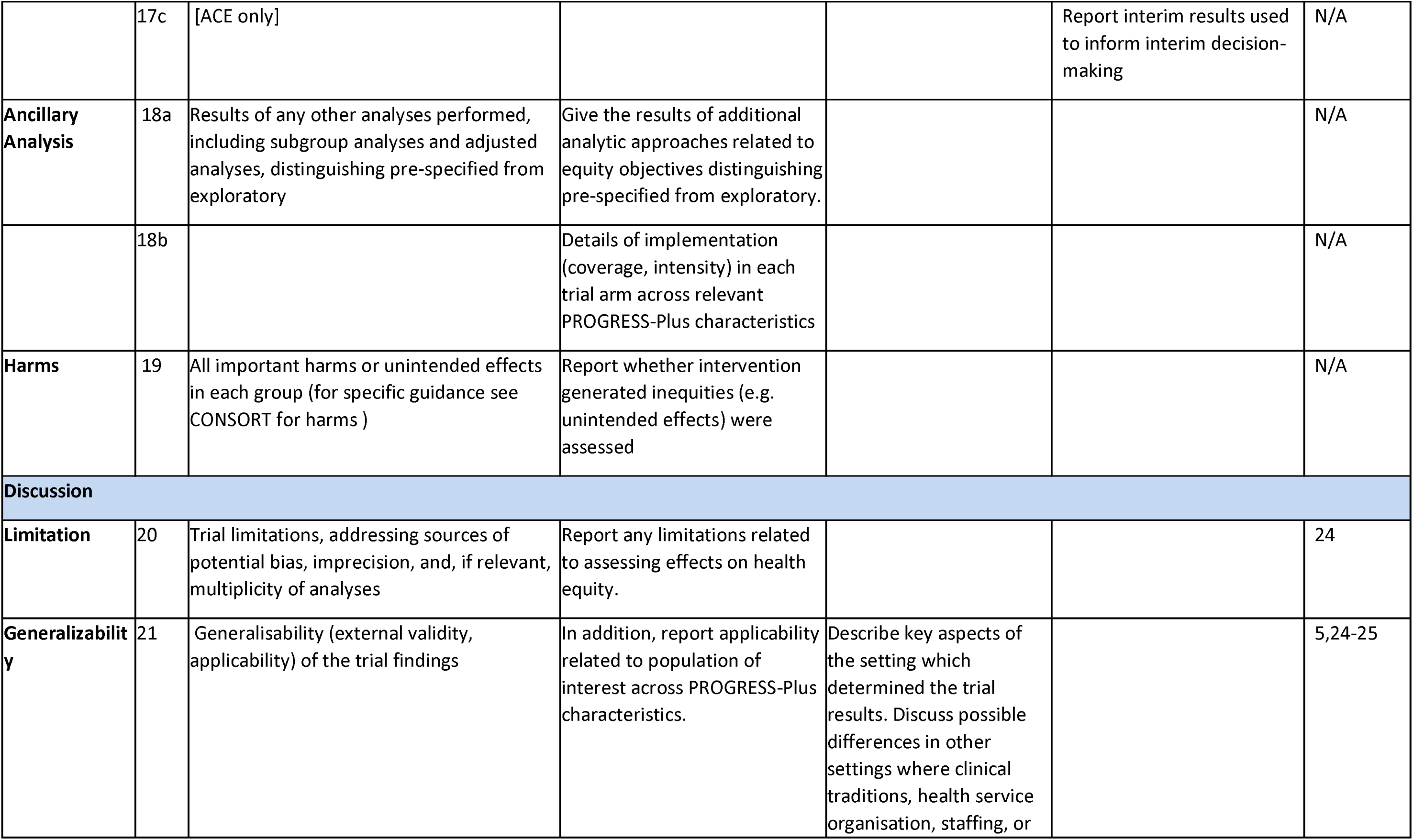
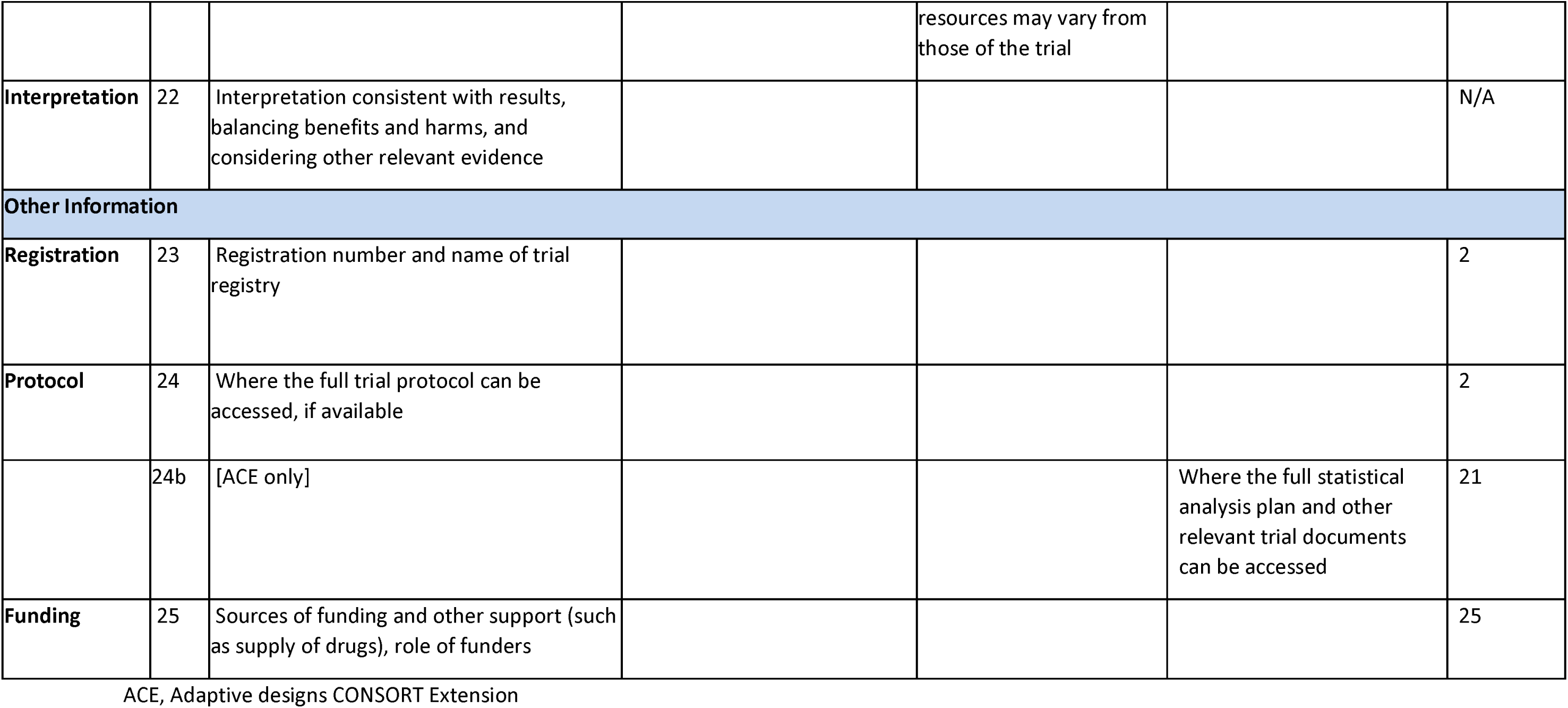

## Appendix: Data management plan

### 1. DATA SOURCES AND DATA COLLECTION PROCESSES

The research objectives require the collection of quantitative survey data, as well as qualitative data in the form of audio recordings and quotes from study participants. Table 1 below outlines the data fields to be collected throughout the various stages of the data collection process. All data will be treated as personal data for the purpose of data capturing and processing, as collectively, it can be combined in a way that could make it identifiable.

Data from the initial screening process will be collected in Peek powered Eye Health School and Community Programmes using Peek’s Capture application. During the initial screening process only basic and non-personal identifying data is collected, with the exception of telephone number. Following initial screening, all those identified as requiring referral will be asked to provide sociodemographic data to enable us to monitor the equity performance of our programmes e.g. are certain ethnic groups more likely to be screened? The additional sociodemographic indicators are outlined in table 1 below. Based on the visual acuity threshold set prior to screening, the Peek Capture automatically informs the data collector whether the attendee may potentially need onward treatment. For those screened negative no further data is collected. Only for those screened positive is further information collected. This ensures data collection is kept to an absolute minimum maintaining privacy and ensuring compliance with data protection regulations. For those screened positive, additional information is collected, but the data is always minimised to ensure only the required data is collected at each stage of the service.

Following triage of individuals who had screened positive, a four-stage rapid exploratory sequential mixed-methods study design will be used to evaluate barriers to health access among non-attenders who had been flagged for onward treatment. Telephone interviews will be conducted among 60 non-attenders, purposively selected from socio-demographic groups with the lowest overall attendance rates. The aim of the telephone interviews is to explore and evaluate their perceived barriers to clinic attendance, and develop a list of potential solutions. Once interventions and service modifications have been identified, these will be tested through a series of pragmatic, embedded, adaptive parallel, multi-arm randomized control trials (APT). The intention of the APT is to continuously improve attendance rates, particularly amongst those groups with the lowest engagement rates overall. Table 1 outlines each of the data collection phases, the data fields to be collected, and the study populations of each of the stages discussed.

**Table 1:**
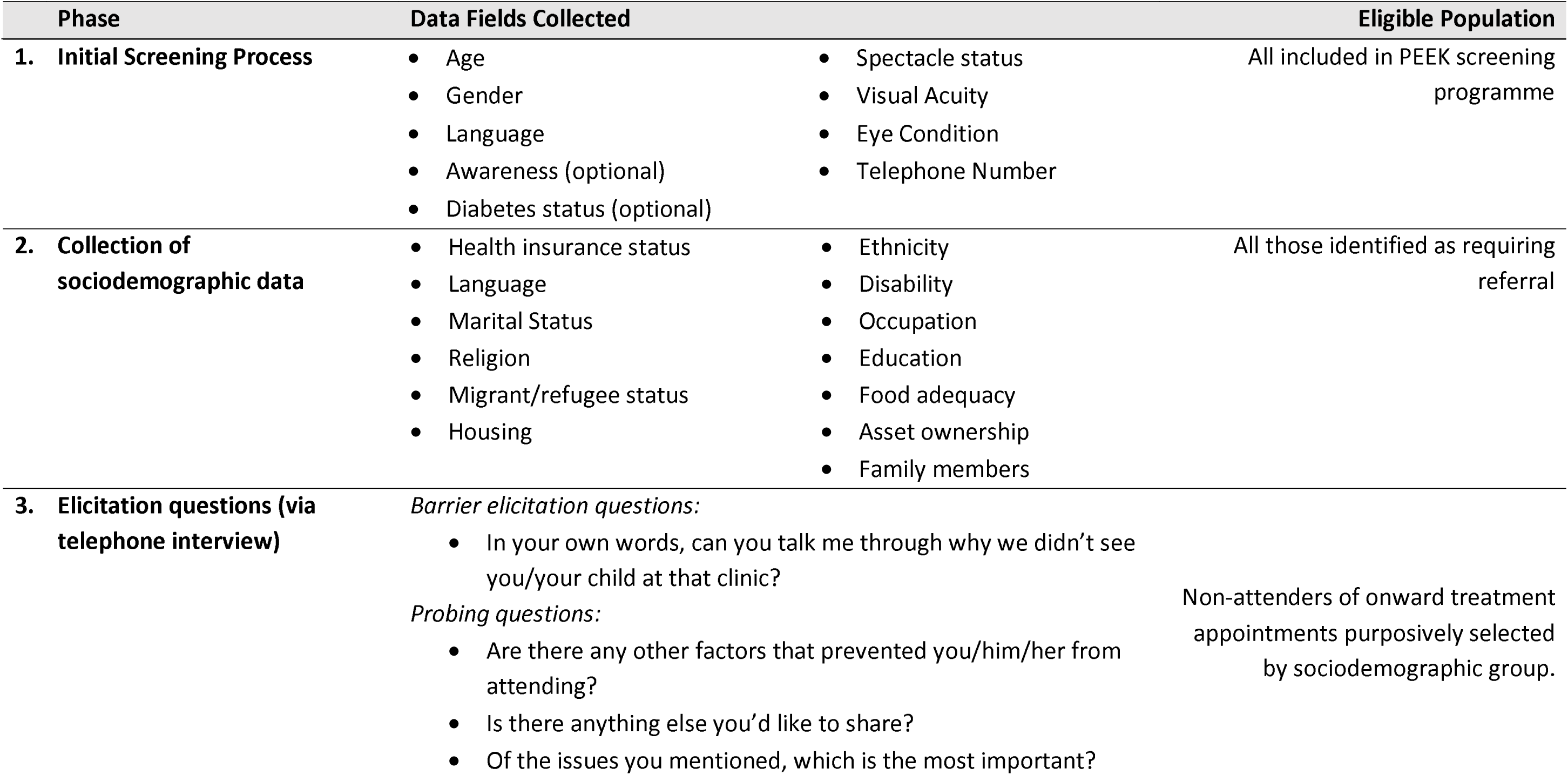
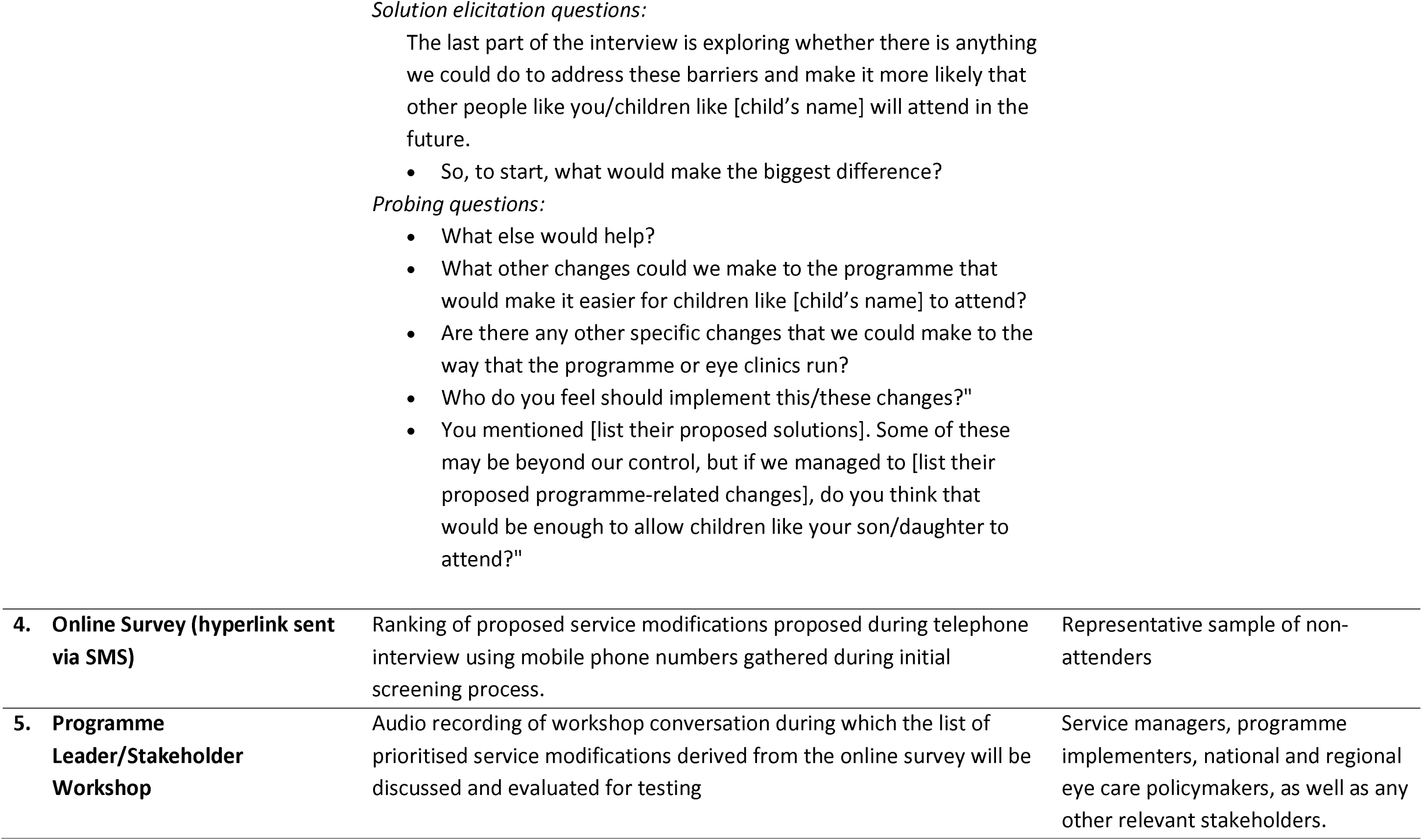
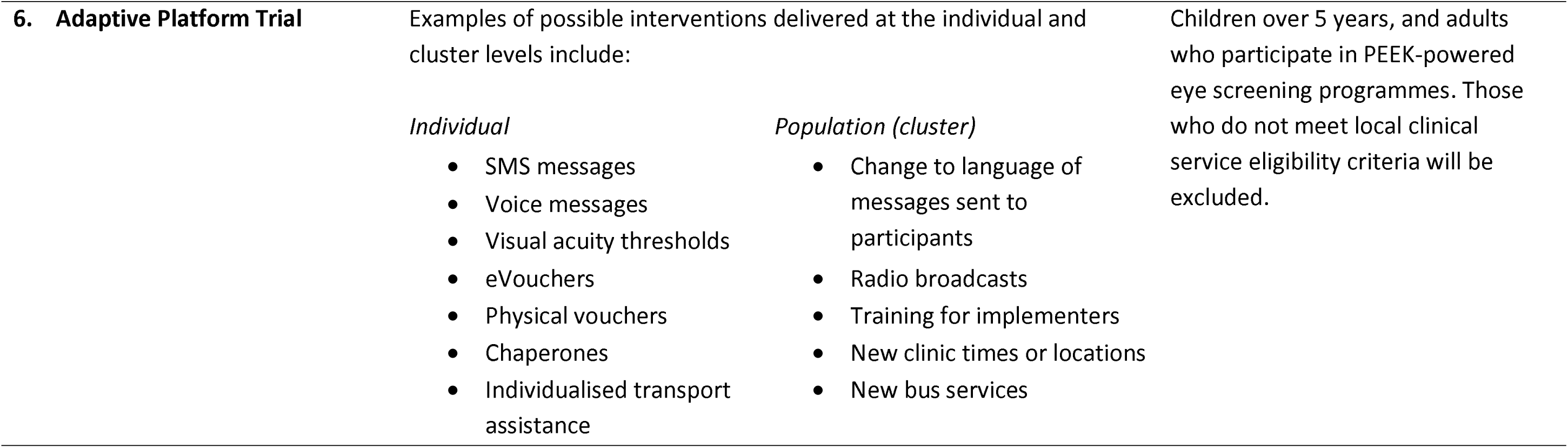
Data collection phases, data fields and study populations for broader I’M SEEN project

### 2. DATA COLLECTION TOOLS

#### Various data collection tools will be used to populate the data fields outlined in table 1. Quantitative Data

- <u>Android Mobile Devices</u> – Survey data, and data derived from the APT (phases 1,2, 4 and 6) will be collected by Peek’s implementing partners using Android devices through the Peek Capture application. Peek Capture enforces security controls that include strong device passcodes and native Android encryption. Data stored is time limited, the device syncs via an encrypted connection with a Peek managed server, the data is then deleted to minimise the risk of data stored on the device. The APT will be embedded within Peek software used in parallel with a Bayesian algorithm that will be used to autonomously run response adaptive trials.

#### Qualitative Data

- Play Verto – The online survey will be administered through Play Verto, a play-based online survey group who have worked with the United Nations and others to develop engaging short surveys that have impressively high response rates in low- and middle-income countries. The survey will be sent as a hyperlink in an SMS. PlayVerto will gather, store and process. After, they will transfer (anonymised data) it to LSHTM who will perform further processing and storage. LSHTM will share aggregate anonymised findings with partners and in public domain.
- Data Abstraction Matrix: During the telephone interviews, data collectors will directly enter notes, quotes, open codes, and abstractions into a matrix. Data gathered, processed and stored by local partner organization. Then shared with LSHTM (fully-anonymised responses to be shared).
- Audio Recordings – Telephone interviews will involve verbal communication and discussion, and thus will be collected and stored using digital audio-recording methods.

#### Software

- Peek Capture - is an application that runs on Android devices that supports eye health screening and referral pathways to treatment
- Peek Admin - is a web based data platform application that is used to view the data collected by Peek Capture, it tracks the Programme progress, provides insights and helps ensure no one is left behind.
- Play Verto – is a play-based online survey group who have worked with the United Nations and others to develop engaging short surveys that have impressively high response rates in low- and middle-income countries.
- STATA and R, and Excel will be used to analyse the data exported from Peek Admin

#### Hardware

- Peek servers are hosted on Amazon Elastic Compute cloud-based virtual machines running Amazon Linux.
- Android devices, locally managed by Peek’s implementing partners.

### 3. DATA-RELATED ACTIVITIES

| Task | Description |
| --- | --- |
| Start gathering SES data | In month 1 we will start gathering sociodemographic data from:<br>• a representative sample of all those presenting to be screened<br>• all those identified with an eye care needs and referred on for treatment<br>These data will be transferred from Android devices in the field to Peek Admin, hosted on AWS.<br>Note that Peek programmes run continuously and we intend to gather data from participants in every programme so that we can promote equitable service delivery. |
| Clean SES data | Routine manual data cleaning will be conducted periodically by Peek administrators. Internal software guardrails will pick up simple errors |
| Analyse SES data | Every month we will perform simple descriptive statistical analysis of presentation rates and treatment attendance rates by SES category.<br>The output of this analysis will be anonymised and presented as mean attendance rates for each SES subgroup e.g. males x%, females z%. |
| Conduct telephone interviews, online surveys and stakeholder workshop | In order to better understand barriers to accessing eye services a series of activities will be conducted through a four-stage sequential mixed-methods approach. These include:<br>1. Telephone Interviews – Telephone interviews will be conducted with non-attenders, purposively selected from subgroups with the lowest attendance rates.<br>2. Following telephone interviews, a single list of suggested solutions will be compiled<br>3. Online survey – An online survey will be conducted among a representative sample of non-attenders to rank mooted interventions/service modifications.<br>4. Stakeholder workshop – Programme leaders and key stakeholders will then select one or more of the highest ranked interventions to test, based on impact, feasibility, risk and cost. |
|  | Following completion of this process, data will be analysed to elicit barriers to care and recommended interventions/service modifications to improve attendance rates. |
| Testing of service modifications through APT | An automated adaptive platform trial (APT) will iteratively test a series of interventions selected with intended service beneficiaries to increase attendance rates among marginalised groups. This will be done through a Bayesian, embedded, pragmatic, superiority, adaptive platform trial platform that will use response adaptive randomisation. |

#### Quality checks

- Errors are flagged at the point of data entry by software that only accepts pre-specified responses e.g. phone numbers must be comprised of a set string length of digits.
- The software has built-in logic steps
- We will institute training and supervision for all data collectors
- Application logging, audit trails and alerting direct administrators to given issues post-collection e.g. when SMS messages fail to be delivered
- Post-collection human data checking using the Peek Admin programme e.g. for ID disambiguation

5. How will you address ethical & legal issues within your research?

- What permissions are needed? E.g. to collect data in country, analyse data for specific purpose, share data
- From whom must approval be obtained? E.g. study participant, ethics committees, data provider
- How will permissions be provided? E.g. ask participants to sign a consent form, sign a Data Transfer Agreement

### 4. PERMISSIONS

Local permissions for Peek powered eye health programmes are already in place. This is in the form of data processing agreements with Peek and the local MoH and/or local implementing partner. This provides a legal agreement between the parties that the data can be collected and processed. The proposed research will be authorised by the same parties to ensure full transparency and the data collection and processing will be managed under the same data processing agreement.

We will obtain written informed consent to collect, analyse, and publish anonymised aggregate participant data in peer-reviewed journals and online open-access data repositories. Individuals will not be identifiable.

In line with UK guidance on risk-adapted approaches to obtaining informed consent, participants will provide consent by ticking a box underneath the following statement:

> *“I understand that my anonymous data may be shared with other researchers or online, and that I will not be identifiable from this information. I understand that my decision will not affect the care that I receive, and I am free to change my mind anytime I like.”*

Consent will be obtained when participants initially present for screening.

For screening programmes that include children (<18 years), we will seek consent from their parents/legal guardians using the following statement, sent home on a paper form along with the generic participant information leaflets before screeners visit the school:

> *“I understand that my child’s anonymous data may be shared with other researchers. I understand that my child will not be identifiable from this information. I understand that my decision will not affect the care that my child receives, and I am free to change my mind anytime I like.”*

Approval will be sought from research ethics committees at LSHTM and each of the countries where screening takes place.

### 5. DOCUMENTATION

Standard operating procedures and an overall study protocol will be developed in line with LSHTM research guidance to cover all aspects of the research project.

Standardised online training modules have been delivered for programme implementing partners tasked with data collection in the field.

Training will be delivered to all project staff to ensure that they understand the requirements and are able to follow the SOPs.

We have a data compendium which describes the custom sociodemographic variables that we will collect in each country,

### 6. DATA STORAGE AND SECURITY

Data collection, management and storage for this study will be managed by seven entities described below:

A. Peek Vision Capture Application
B. Play Verto
C. The London School of Hygiene and Tropical Medicine
D. Botswana: The University of Botswana
E. India: Dr Shroff Charity Eye Hospital
F. Kenya: Kenya Medical Research Institute?
G. Nepal: Nepal Netra Jyoti Sangh

#### Peek Capture Application

##### Pre research data collection and storage in Peek powered eye health programmes

The data will be collected in Peek powered Eye Health School and Community Programmes using Peek’s Capture application. Data will be collected by Peek’s implementing partners using Android devices through the Peek Capture application. Peek Capture enforces security controls that include strong device passcodes and native Android encryption. Data stored is time limited, the device syncs via an encrypted connection with a Peek managed server, the data is then deleted to minimise the risk of data stored on the device. h

The data is stored on a Peek managed server hosted in a Virtual Private Cloud (VPC) utilising the Amazon Web Services (AWS) Cloud. Each Peek powered programme is hosted on it’s own dedicated server and a VPC that will reside in the UK/EU ensuring all of the data privacy safeguards as governed under the GDPR. All data collected is securely stored in AWS data centers which are state of the art, utilising innovative architectural and engineering approaches. More information, including a virtual tour, can be found by visiting the link <u>here</u>.

Throughout the eye health programme life cycle only approved implementation partners and Peek team members have access to programme data. Access is strictly controlled through the Peek Admin web based data platform application. This is used to view the data collected by Peek Capture, it tracks the Programme progress, provides insights and helps ensure no one is left behind.

##### Peek Capture security

- Peek Capture is installed on implementing partners managed Android devices
- Peek Capture enforces security controls that include strong device passcodes and native Android encryption.
- Data stored is time limited, the device syncs via an encrypted connection with a Peek managed server, the data is then deleted to minimise the risk of data stored on the device.

##### Peek Admin security

- Strong passwords, minimum of 12 characters, password strength meter where only ‘strong’ is accepted, blacklist passwords are enforced to ensure easily guessed and passwords found in data breaches cannot be used.
- 2-Factor Authentication to protect user account security.
- User access permissions are controlled through account privileges, this controls scope of programme so access is restricted and limited to only what a user requires for their work, admin privileges are restricted to only those that require the access, account management and patient level reporting.
- Accounts disable automatically after 60 days of inactivity.
- User access reviews available for implementing partners to ensure leavers and inactive accounts are removed.

##### Peek Platform Data Security Assurance

Peek is an International Standardisation Organisation (ISO) 27001 certified organisation. ISO 27001 certification requires an annual audit by an accredited external auditing body who verify compliance with the industry best practice information security controls.

Peek servers hosted in a Virtual Private Cloud (VPC) utilising the Amazon Web Services (AWS) Cloud. Each Peek powered programme is hosted on it’s own dedicated server and a VPC that will reside in the UK/EU ensuring all of the data privacy safeguards as governed under the GDPR. All data collected is securely stored in AWS data centers which are state of the art, utilising innovative architectural and engineering approaches.

More information, including a virtual tour, can be found by visiting the link below: https://aws.amazon.com/compliance/data-center/.

Annual penetration tests conducted by a 3rd party specialist security testing company. The purpose of the test is to verify whether robust security mechanisms are in place to prevent unauthorised users from accessing data and infrastructure. This penetration test includes:

- Identification of potential vulnerabilities occurring in the application and defining possible attack scenarios conducted with techniques typical for attacks on web applications;
- Simulated attacks from the perspective of an anonymous and standard user;
- Testing API endpoints from the perspective of an anonymous and standard user, including mechanisms such as user authentication, access control, and data validation;
- Security assessment of our infrastructure against the latest industry standard AWS CIS Foundations Benchmark.

The AWS Compliance Program provides further assurance and understanding of the robust controls in place to maintain security and compliance in the cloud. AWS regularly achieves third-party validation for thousands of global compliance requirements that are continuously monitored to meet security and compliance standards for the most sensitive data and privacy requirements. AWS supports more security standards and compliance certifications than any other offering, including PCI-DSS, HIPAA/HITECH, FedRAMP, GDPR, FIPS 140-2, and NIST 800-171, helping satisfy compliance requirements for virtually every regulatory agency around the globe. More information can be found by visiting https://aws.amazon.com/compliance/programs/.

##### Peek Platform Data Security Controls

###### Peek Servers

Peek servers hosted in a Virtual Private Cloud (VPC) utilising the Amazon Web Services (AWS) Cloud. Each Peek powered programme is hosted on it’s own dedicated server and a VPC that will reside in the UK/EU ensuring all of the data privacy safeguards as governed under the GDPR.

Server OS is Amazon Linux ustlising AWS AMIS to provide base images for our system drives and enhances security by focusing on two main security goals, limiting access and reducing software vulnerabilities. Security updates are applied automatically to test once a week and then rolled out a week later automatically to other environments

###### Docker

Peek server software runs in Docker containers. Docker shields application software from variations in platform and co-hosted software. It ensures that development, test and production environments run the same context as one another to ensure consistent, predictable behaviour. Peek servers also use docker swarm mode to achieve failsafe reliability and replication of Mongo databases.

###### Databases

Server data is stored in Mongo databases, a fast, scalable, json document database. Peek infrastructure uses a Mongo replica set across two hosts. There are two replicas each holding a full copy of the data and one arbiter. The arbiter is only used for the election of a new master if one of the nodes was to become unavailable. The Mongo database and journal are held on AWS Secure EBS volumes. This provides 256-bit AES encrypted using a key managed under the Amazon Key Management Service.

Amazon Key Management Service, allows us to create and manage cryptographic keys and securely control their use across a wide range of AWS services and within our applications. AWS KMS is a secure and resilient service that uses hardware security modules that have been validated under FIPS 140-2 to protect the encryption keys. AWS KMS also integrates with AWS CloudTrail providing us with secure logs of all key usage. Backups on S3 are also encrypted using keys managed by AWS Key Management Service.

###### Logging and Monitoring

Peek Server and Mongo Server logs and uploaded to AWS Cloudwatch for storage and monitoring. AWS Cloudwatch collects monitoring and operational data in the form of logs, metrics, and events and alerts us immediately of problems in any environment, both application and infrastructure.

###### Network Security

AWS Security groups are used to provide firewall-like network access control and allow inbound traffic on HTTP and HTTPS ports. Outbound traffic is permitted on any port. The SSH traffic is restricted to subnets associated with devops engineers and the deployment servers. TLS 1.2 is used to secure traffic between device or browser and server.

Operational access to the AWS console is protected with AWS IAM MFA which uses 2-Factor Authentication and ensures that access to AWS is restricted to users with knowledge of password and possession of a specific approved mobile device. Automated access to the AWS API uses AWS Roles with restricted privileges needed for housekeeping, logging and alarm maintenance. No user use is made of Access Keys to eliminate the vulnerabilities of file-system-based credentials.

###### Threat Detection

AWS Guard Duty is enabled, this provides a threat detection service that continuously monitors for malicious activity and unauthorised behaviour to protect access, workloads and data. The service utilises up-to-date threat intelligence feeds from AWS, CrowdStrike, and Proofpoint and continuously evolves through machine learning.

###### Backups

An Image is maintained of the Server Host using AWS AMI to ensure continuous availability.

A snapshot of the encrypted data volume, containing database and journal, is taken four times daily. Snapshots are retained for two weeks. Access to the snapshots is strictly controlled. Old backups are automatically deleted after 90 days. Backups are stored on AWS S3 storage, also encrypted providing 256-bit AES encryption. The backups are stored across AWS multiple availability zones, this ensures that the data resides in multiple data centres separated geographically and stored in AWS secure data centres.

Additionally, a further backup is made off AWS. Off-AWS backups are replicated to Google Cloud daily via Google Transfer service to identically named buckets and files with a retention policy of 90 days.

###### Data Centres

All data collected is securely stored in AWS data centers which are state of the art, utilising innovative architectural and engineering approaches.

###### Disaster Recovery

A full disaster recovery test is performed at least annually to ensure servers, applications and databases can be fully recovered within 24 hours.

##### Play Verto

###### Play Verto Data capture tool

Data collection via our web-based application is all stored on a AWS RDS dedicated server, located in Ireland. This database utilises AWSs own encryption, AES-256 at rest, for maximum security. All data collected is securely stored in AWS data centers which are state of the art, utilising innovative architectural and engineering approaches. More information, including a virtual tour, can be found by visiting the link <u>here</u>.

Only approved team members have access to the data. Access is strictly controlled through the Play Verto’s Admin and AWS Admin. Where Password protection is required and the use of 2-factor authentication where applicable.

###### Play Verto Capture security

- Play Verto is a web-based application therefore can only be accessed via a public URL.
- Play Verto enforces security controls that include strong device passcodes and 2-factor authentication where applicable…
- Data stored is encrypted via AES-256 encryption

###### Play Verto Admin security

- We have a strong password policy in place for all our accounts, requiring a minimum length of 8 characters.
- 2-Factor Authentication to protect user account security.
- User access permissions are controlled through account privileges. So access is restricted and limited to only what a user requires for their work.

###### Play Verto Platform Data Security Assurance

Play Verto complies with CyberEssentials Certification and IASME Governance Standard. Data collection via our web-based application is all stored on a AWS RDS dedicated server, located in Ireland. This database utilises AWSs own encryption, AES-256 at rest.

Monthly automated penetration tests conducted by <u>Detectify</u> The purpose of the test is to verify whether robust security mechanisms are in place to prevent unauthorised users from accessing data and infrastructure. We have maintain Threat score of 0 and 10/10, OSWASP SCORE (The worldwide non-profit organization Open Web Application Security Project (OWASP)’s list of the ten most common vulnerabilities, known as OWASP Top 10, is often used as a security standard. Detectify covers OWASP Top 10 and provides an easy way for you to see which categories you pass or fail.)

The AWS Compliance Program provides further assurance and understanding of the robust controls in place to maintain security and compliance in the cloud. AWS regularly achieves third-party validation for thousands of global compliance requirements that are continuously monitored to meet security and compliance standards for the most sensitive data and privacy requirements. AWS supports more security standards and compliance certifications than any other offering, including PCI-DSS, HIPAA/HITECH, FedRAMP, GDPR, FIPS 140-2, and NIST 800-171, helping satisfy compliance requirements for virtually every regulatory agency around the globe. More information can be found by visiting https://aws.amazon.com/compliance/programs/.

##### Play Verto Platform Data Security Controls

###### Play Verto Servers

Data collection via our web-based application is all stored on a AWS RDS dedicated server, located in Ireland. This database utilises AWSs own encryption, AES-256 at rest, for maximum security. Ensuring all of the data privacy safeguards as governed under the GDPR.

###### Databases

Server data is stored in Mongo databases, a fast, scalable, json document database. Play Verto infrastructure uses a Mongo replica set across two hosts. There are two replicas each holding a full copy of the data and one arbiter. The arbiter is only used for the election of a new master if one of the nodes was to become unavailable. The Mongo database and journal are held on AWS Secure EBS volumes. This provides 256-bit AES encrypted using a key managed under the Amazon Key Management Service.

Amazon Key Management Service, allows us to create and manage cryptographic keys and securely control their use across a wide range of AWS services and within our applications. AWS KMS is a secure and resilient service that uses hardware security modules that have been validated under FIPS 140-2 to protect the encryption keys. AWS KMS also integrates with AWS CloudTrail providing us with secure logs of all key usage. Backups on S3 are also encrypted using keys managed by AWS Key Management Service.

###### Logging and Monitoring

Play Verto Server and Mongo Server logs and uploaded to AWS Cloudwatch for storage and monitoring. AWS Cloudwatch collects monitoring and operational data in the form of logs, metrics, and events and alerts us immediately of problems in any environment, both application and infrastructure.

###### Network Security

AWS Security groups are used to provide firewall-like network access control and allow inbound traffic on HTTP and HTTPS ports. Outbound traffic is permitted on any port. The SSH traffic is restricted to subnets associated with devops engineers and the deployment servers. TLS 1.2 is used to secure traffic between device or browser and server.

Operational access to the AWS console is protected with AWS IAM MFA which uses 2-Factor Authentication and ensures that access to AWS is restricted to users with knowledge of password and possession of a specific approved mobile device. Automated access to the AWS API uses AWS Roles with restricted privileges needed for housekeeping, logging and alarm maintenance. No user use is made of Access Keys to eliminate the vulnerabilities of file-system-based credentials.

###### Threat Detection

AWS Guard Duty is enabled, this provides a threat detection service that continuously monitors for malicious activity and unauthorised behaviour to protect access, workloads and data. The service utilises up-to-date threat intelligence feeds from AWS, CrowdStrike, and Proofpoint and continuously evolves through machine learning.

###### Backups

An Image is maintained of the Server Host using AWS AMI to ensure continuous availability.

A snapshot of the encrypted data volume, containing database and journal, is taken four times daily. Snapshots are retained for two weeks. Access to the snapshots is strictly controlled. Old backups are automatically deleted after 90 days. Backups are stored on AWS S3 storage, also encrypted providing 256-bit AES encryption. The backups are stored across AWS multiple availability zones, this ensures that the data resides in multiple data centres separated geographically and stored in AWS secure data centres.

Additionally, a further backup is made off AWS. Off-AWS backups are replicated to Google Cloud daily via Google Transfer service to identically named buckets and files with a retention policy of 90 days.

###### Data Centres

All data collected is securely stored in AWS data centres which are state of the art, utilising innovative architectural and engineering approaches.

###### Disaster Recovery

A full disaster recovery test is performed at least annually to ensure servers, applications and databases can be fully recovered within 24 hours.

## EXPORT DATA SHARING FOR ANALYSIS

At the analysis stage pseudo-anonymised data will be exported in an encrypted zip file CSV file to LSHTM researchers to perform statistical testing. The zip file will be saved on the protected LSHTM server and only named project staff will be given access. Passwords will be sent separately. We will only ever export the minimum data required for the analyses.

### Labelling conventions

1. Keep file names short, meaningful and easily understandable to others.
2. Order the elements in a file name in the most appropriate way to retrieve the record.
3. Avoid unnecessary repetition and redundancy in file names and paths
4. Avoid obscure abbreviations and acronyms. Use agreed University abbreviations and codes where relevant.
5. Avoid vague, unhelpful terms such as “miscellaneous” or “general” or “my files”
6. Use capital letters to delimit words, as the preferred option, although underscores (_) or hyphens (-) may add clarity, they make the file name longer.
7. For numbers 0-9, always use a minimum of two digit numbers to ensure correct numerical order (e.g. 01, 02, 03 etc.)
8. Dates should always follow same format: YYYY-MM-DD e.g. 2017-04-25
9. When including a personal name give the family name first followed by initials, with no comma in between e.g. SmithAB
10. Avoid using common words such as ‘draft’ or ‘letter’ at the start of file names unless doing so will make it easier to retrieve the record.
11. Use alphanumeric characters i.e. letters (A-Z) and numbers (0-9). Avoid using invalid characters in file names such as *? \ / : # % ∼ { }
12. The file names of records relating to recurring events should include the date and a description of the event, except where the inclusion of these elements would be incompatible with rule 3.
13. The version number of a record should be indicated in its file name by the inclusion of ‘V’ followed by the version number (e.g. V01, V03 etc.). However versioning is enabled automatically in systems such as Office 365 and One Drive for Business, making it unnecessary to duplicate this information in the file name itself. e.g. 2021-11-19_Topic_Filename-variable01

How will we keep data safe and secure?

- Delete personal & confidential details at the earliest opportunity (specify when)
- Use digital storage that require a username/password or other security feature
- Physical security (such as locked cabinet or room)
- Encrypt storage devices
- Encrypt data during transfer
- Avoid cloud services located outside EU
- Take ‘Information Security Awareness training’
- Ensure backups are also held securely

The aggregated data that is shared among project staff and partners will not contain any names, however the data being shared may still permit the identification of individuals depending on the domains being shared and may therefore constitute pseudo-anonymised data.

We also note that there is not adequate shared secure storage space at LSHTM. We will have to use our personal H drives which is suboptimal for joint working and version control.

## ARCHIVING & SHARING

All data will be stored for 10 years.

- Files intended for sharing may be hosted in the LSHTM data repository (http://datacompass.lshtm.ac.uk) or a 3rd party repository, such as UK Data Service, ArrayExpress, Zenodo, etc.
- Internal and confidential files can be held on the LSHTM Secure Server
- Internal confidential files will be retained on Peek’s secure servers.
- LSHTM analyses will be saved on encrypted and password-protected files on LSHTM SharePoint, with access restricted to the project team. Once the project is complete these files will be moved to a secure server.
- Data presented in publications (anonymised aggregate mean attendance rates for each SES subgroup) will be published on GitHub.

Resources will be made available at the same time as findings are published in an academic journal. Once available, we will make other researchers aware that the resources exist by:

- Citing resources in future research papers, e.g. in the data access statement or reference list
- Citing resources in project reports
- Adding resources to a list of our academic outputs

The following steps will be taken to ensure that resources are easy to analyse and use in future research:

- Store resources in open file formats such as CSV, Rich Text, etc. See https://www.ukdataservice.ac.uk/manage-data/format/recommended-formats
- Designate a corresponding author / data custodian who will handle data-related questions

### Conditions on access/use

| Requirement: | To be addressed by: |
| --- | --- |
| In line with the UK concordat on open research data (2016), anonymised data from this trial will be made available to bona fide research groups (evidenced via CVs and the involvement of a qualified statistician), and in line with the trial's publicly available data sharing policy, following review and approval from the trial's data monitoring committee. No reasonable request will be turned down, and the appropriate data will be made available within 1-month of receiving the request. | The PI will forward requests for data to the in-country leads in order to seek the relevant permissions. We will respond to any bona fide request within 28 days. |
| There may be multiple levels of permission required in-country before data can be shared, including national ministry of health approval and local implementation partner approval |  |

## RESOURCING

With respect to costs of resources, we have adequate funding within the Wellcome project grant. The data is collected through active live Peek powered programmes where funding and resources is already provided for data collection and data security.

