## Supplementary material for "Protocol for an adaptive platform trial of intended service user-derived interventions to equitably reduce non-attendance in eye screening programmes in Botswana, India, Kenya & Nepal": CONSORT Statement

Appendix: CONSORT checklists

| **Section** | **Item** | **Standard 2010 CONSORT Item** | **Equity extension** | **Pragmatic extension** | **Adaptive extension** | **Page** |
| --- | --- | --- | --- | --- | --- | --- |
| **Title** | | | | | | |
| **Title** | 1a | Identification as a randomised trial in the title | If health equity is a major focus, consider using the term “health equity” in the title. |  |  | 1 |
| **Abstract** | | | | | | |
| **Structured Summary** | 1b | Structured summary of trial design, methods, results, and conclusions (for specific guidance see CONSORT for abstracts) | State research question(s) related to health equity |  |  | 2 |
|  | 1c |  | Present results of all planned health equity analyses |  |  | 2 |
|  | 1d |  | Describe extent and limits of applicability to populations of interest across PROGRESS-Plus characteristics |  |  | 2 |
| **Introduction** | | | | | | |
| **Background** | 2a | Scientific background and explanation of rationale | Describe rationale for focus on health equity | Describe the health or health service problem that the intervention is intended to address and other interventions that may commonly be aimed at this problem |  | 3,5,6 |
| **Objective** | 2b | Specific objectives or hypotheses | State the objective being addressed with reference to health equity |  |  | 7 |
| **Methods** | | | | | | |
| **Trial Design** | 3a | Description of trial design (such as parallel, factorial) including allocation ratio | Describe aspects of trial design that were chosen to answer equity questions |  |  | 7 |
|  | 3b | Important changes to methods after trial commencement (such as eligibility criteria), with reasons |  |  | Type of adaptive design used, with details of the pre-planned trial adaptations and the statistical information informing the adaptations  Important changes to the design or methods after trial commencement (such as eligibility criteria) outside the scope of the pre-planned adaptive design features, with reasons | 12-13 |
| **Participants** | 4a | Eligibility criteria for participants | Report population eligibility criteria across relevant PROGRESS-Plus characteristics. | Eligibility criteria should be explicitly framed to show the degree to which they include typical participants and/or, where applicable, typical providers (eg, nurses), institutions (eg, hospitals), communities (or localities eg, towns) and settings of care (eg, different healthcare financing systems) |  | 8-9, 14 |
|  | 4b | Settings and locations where the data were collected | Report context and relationship to health inequity |  |  | 7 |
|  | 4c |  | Report details of partnerships with populations and communities, where applicable. |  |  | 5,7,23 |
| **Intervention** | 5 | The interventions for each group with sufficient details to allow replication, including how and when they were actually administered | Report whether comparator intervention is the standard of care, and whether it has equity implications. | Describe extra resources added to (or resources removed from) usual settings in order to implement intervention. Indicate if efforts were made to standardise the intervention or if the intervention and its delivery were allowed to vary between participants, practitioners, or study sites  Describe the comparator in similar detail to the intervention |  | 8,11-12 |
| **Outcomes** | 6a | Completely defined pre-specified primary and secondary outcome measures, including how and when they were assessed | Report whether outcomes were identified as relevant and important to population(s) across PROGRESS-Plus characteristics and how this was done | Explain why the chosen outcomes and, when relevant, the length of follow-up are considered important to those who will use the results of the trial | Any other outcome measures used to inform pre-planned adaptations should be described with the rationale | 13 |
|  | 6b | Any changes to trial outcomes after the trial commenced, with reasons |  |  |  | N/A |
| **Sample Size** | 7a | How sample size was determined | Report whether analyses focused on health equity objectives are powered to detect differences. | If calculated using the smallest difference considered important by the target decision maker audience (the minimally important difference) then report where this difference was obtained | How sample size and operating characteristics were determined | 15-16 |
|  | 7b | When applicable, explanation of any interim analyses and stopping guidelines |  |  | Pre-planned interim decision-making criteria to guide the trial adaptation process; whether decision-making criteria were binding or non-binding; pre-planned and actual timing and frequency of interim data looks to inform trial adaptations | 15-18 |
| **Randomisation Sequence Generation** | 8a | Method used to generate the random allocation sequence |  |  |  | 19 |
|  | 8b | Type of randomisation; details of any restriction (such as blocking and block size) | Report whether randomisation was stratified on PROGRESS-Plus characteristic(s) |  | Type of randomisation; details of any restriction (such as blocking and block size); any changes to the allocation rule after trial adaptation decisions; any pre-planned allocation rule or algorithm to update randomisation with timing and frequency of updates | 19 |
| **Allocation Concealment Mechanism** | 9 | Mechanism used to implement the random allocation sequence (such as sequentially numbered containers), describing any steps taken to conceal the sequence until interventions were assigned |  |  |  | 19 |
| **Implementation** | 10 | Who generated the random allocation sequence, who enrolled participants, and who assigned participants to interventions |  |  |  | 19 |
| **Blinding** | 11a | If done, who was blinded after assignment to interventions (for example, participants, care providers, those assessing outcomes) and how |  | If blinding was not done, or was not possible, explain why |  | 20 |
|  | 11b | If relevant, description of the similarity of interventions |  |  |  | 11-12, 19-20 |
|  | 11c | [only applies for ACE] |  |  | Measures to safeguard the confidentiality of interim information and minimise potential operational bias during the trial | 20-22 |
| **Statistical Methods** | 12a | Statistical methods used to compare groups for primary and secondary outcomes |  |  | Statistical methods used to compare groups for primary and secondary outcomes, and any other outcomes used to make pre-planned adaptations | 15-18, 21 |
|  | 12b | Methods for additional analyses, such as subgroup analyses and adjusted analyses | Report details of additional analyses focused on health equity, including whether analyses to estimate heterogeneity of effects between population subgroups were done on an additive or multiplicative scale, and whether pre-specified. |  |  | 16, 21 |
|  | 12c | [ACE only] |  |  | For the implemented adaptive design features, statistical methods used to estimate treatment effects for key endpoints and to make inferences | 21 |
| **Ethical Concerns** | a | [equity only] | Report details of ethical clearance and informed consent |  |  | 19,23 |
| **Results** | | | | | | |
| **Participant flow (a diagram is strongly recommended)** | 13a | For each group, the numbers of participants who were randomly assigned, received intended treatment, and were analyzed for the primary outcome | Describe for each group, numbers of participants who were assigned, received and who were analyzed across relevant PROGRESS-Plus characteristics | The number of participants or units approached to take part in the trial, the number which were eligible, and reasons for non-participation should be reported | For each group, the numbers of participants who were randomly assigned, received intended treatment, and were analysed for the primary outcome and any other outcomes used to inform pre-planned adaptations, if applicable | 8,9,15,16 |
|  | 13b | For each group, losses and exclusions after randomisation, together with reasons | Describe for each group, losses and exclusions after randomisation across relevant PROGRESS-Plus characteristics, with reasons. |  |  | 8 |
| **Recruitment** | 14a | Dates defining the periods of recruitment and follow-up | Report whether methods of recruitment were designed to reach populations across relevant PROGRESS-Plus characteristics. |  | Dates defining the periods of recruitment and follow-up, for each group | 5 |
|  | 14b | Why the trial ended or was stopped |  |  |  | 10 |
|  | 14c | [ACE only] |  |  | Specify what trial adaptation decisions were made in light of the pre-planned decision-making criteria and observed accrued data |  |
| **Baseline Data** | 15 | A table showing baseline demographic and clinical characteristics for each group | Present the baseline characteristics also across relevant PROGRESS-Plus characteristics. |  |  | N/A |
|  | 15b | [ACE only] |  |  | Summary of data to enable the assessment of similarity in the trial population between interim stages | N/A |
| **Numbers Analyzed** | 16 | For each group, number of participants (denominator) included in each analysis and whether the analysis was by original assigned groups |  |  |  | N/A |
| **Outcomes and Estimation** | 17a | For each primary and secondary outcome, results for each group, and the estimated effect size and its precision (such as 95% confidence interval) |  |  |  | N/A |
|  | 17b | For binary outcomes, presentation of both absolute and relative effect sizes is recommended |  |  |  | N/A |
|  | 17c | [ACE only] |  |  | Report interim results used to inform interim decision-making | N/A |
| **Ancillary Analysis** | 18a | Results of any other analyses performed, including subgroup analyses and adjusted analyses, distinguishing pre-specified from exploratory | Give the results of additional analytic approaches related to equity objectives distinguishing pre-specified from exploratory. |  |  | N/A |
|  | 18b |  | Details of implementation (coverage, intensity) in each trial arm across relevant PROGRESS-Plus characteristics |  |  | N/A |
| **Harms** | 19 | All important harms or unintended effects in each group (for specific guidance see CONSORT for harms ) | Report whether intervention generated inequities (e.g. unintended effects) were assessed |  |  | N/A |
| **Discussion** | | | | | | |
| **Limitation** | 20 | Trial limitations, addressing sources of potential bias, imprecision, and, if relevant, multiplicity of analyses | Report any limitations related to assessing effects on health equity. |  |  | 24 |
| **Generalizability** | 21 | Generalisability (external validity, applicability) of the trial findings | In addition, report applicability related to population of interest across PROGRESS-Plus characteristics. | Describe key aspects of the setting which determined the trial results. Discuss possible differences in other settings where clinical traditions, health service organisation, staffing, or resources may vary from those of the trial |  | 5,24-25 |
| **Interpretation** | 22 | Interpretation consistent with results, balancing benefits and harms, and considering other relevant evidence |  |  |  | N/A |
| **Other Information** | | | | | | |
| **Registration** | 23 | Registration number and name of trial registry |  |  |  | 2 |
| **Protocol** | 24 | Where the full trial protocol can be accessed, if available |  |  |  | 2 |
|  | 24b | [ACE only] |  |  | Where the full statistical analysis plan and other relevant trial documents can be accessed | 21 |
| **Funding** | 25 | Sources of funding and other support (such as supply of drugs), role of funders |  |  |  | 25 |

ACE, Adaptive designs CONSORT Extension
